# Longitudinal adipose tissue single cell transcriptomics reveals genes and variants regulating weight loss after bariatric surgery

**DOI:** 10.1101/2025.07.11.25331390

**Authors:** Seung Hyuk T. Lee, Asha Kar, Sini Heinonen, Marcus Alvarez, Sandhya Rajkumar, Kristina M. Garske, Kyla Z. Gelev, Birgitta W. van der Kolk, Ulla Säiläkivi, Tuure Saarinen, Dorota Kaminska, Ville Männistö, Markku Laakso, Jussi Pihlajamäki, Anne Juuti, Brunilda Balliu, Kirsi H. Pietiläinen, Päivi Pajukanta

## Abstract

Ability to lose weight during different obesity treatments shows substantial variability between individuals and is likely under genetic control; however, the underlying predictive variants and weight loss genes remain unknown. Here we profiled longitudinal, single cell level adipose transcriptomes of individuals undergoing bariatric surgery to elucidate genes and their regulatory variants contributing to interindividual variability in weight loss outcomes. We identified wide-spread cellular and subcellular transcriptional changes to weight loss with most profound responses in adipocyte subtypes. By clustering the weight loss genes based on their cell-type level co-expression profiles, we uncovered functionally distinct subsets of genes reflecting altered adipocyte expression of central adipocyte function enriched genes. Next, we discovered that body mass index (BMI) polygenic risk score (PRS) built using the *cis* regulatory variants in these 45 adipocyte weight loss genes significantly predict the magnitude of the achieved weight loss and are strongly enriched for variance explained in the change of BMI. Taken together, this longitudinal single nucleus adipose data establishes gene signatures for weight loss and discovers genetic regulators underlying the interindividual variability of weight loss.

## Introduction

The on-going rapid increase in global obesity rates is also increasing the prevalence of extreme obesity, defined as a body mass index (BMI) ≥40 kg/m^2^ ^1^. Individuals with extreme obesity are at a high risk for metabolic dysfunction and multiple cardiometabolic diseases^1^. The propensity towards extreme obesity is partly explained by genetic susceptibility to obesity because individuals with a higher polygenic risk for BMI are more likely to have extreme obesity^2^.

Bariatric surgery is an effective treatment for extreme obesity, resulting in a 40-70% reduction in excess body weight^3^. This also shows that the magnitude of weight loss achieved by bariatric surgery exhibits a substantial interindividual variability, indicating that some individuals benefit more from it than others^4^. However, the cell-type and subcell-type level molecular mechanisms and their genetic contribution to this variability in the weight loss outcome remain elusive.

Subcutaneous adipose tissue (SAT) is a metabolically dynamic fat depot that primarily stores fat by expanding, and thus buffering against obesity^5^. It is also involved in key biological processes, such as energy homeostasis, and serves as an endocrine organ^6^. The capacity to expand allows SAT to act as a buffer against lipotoxicity induced by obesity^5^. Bulk transcriptomic analyses of SAT have shown extensive alterations in gene expression upon change in weight^7^, and recent advances in single cell transcriptomics technologies have revealed highly heterogenous and dynamic nature of SAT cell-types at a single cell resolution, carefully described in the recent adipose atlas^8^. However, little is known about longitudinal cell- and subcell-type level context-specific changes in SAT cell-type abundance and transcription profiles induced by rapid and large weight loss. Furthermore, individual level variability in the SAT cell-types and their contribution to the variance in weight loss outcome is not known due to the lack of longitudinal single cell level data. Similarly, it is not known which particular genes contribute to the weight loss magnitude and to what extent an individual’s achieved weight loss is under genetic control.

In this study, we provide extensive longitudinal single cell level transcriptome profiles of SAT during weight loss by bariatric surgery, integrated with participants’ clinical and genetic data. We identify wide-spread cell-type level changes in SAT expression profiles in response to weight loss, identify cell-type, subcell-type, and context-specific genetic regulation of those weight loss responding genes that contribute to interindividual variability in weight loss outcome; and assess the role of age, sex, and genetic component in the achieved magnitude of the weight loss outcome. Discovering adipocyte weight loss genes and their *cis* regulatory variants allows us to build a response polygenic risk score (PRS) that predicts an individual’s ability to lose weight after bariatric surgery. Overall, we leverage the largest to date longitudinal single nucleus SAT data from four weight loss time-points and demonstrate genetic predisposition to obesity as a significant predictor of weight loss outcome.

## Results

### Generating a longitudinal single nucleus adipose data of weight loss by diet and bariatric surgery

To comprehensively profile cell-type level transcriptional changes due to weight loss, induced by preoperative diet or bariatric surgery, we obtained temporal single nucleus RNA-sequencing (snRNA-seq) data on 167 subcutaneous adipose tissue (SAT) biopsies derived from individuals with grade 3 obesity, who participated in the Roux-en-Y versus one-anastomosis gastric bypass (RYSA) study^9^, a Finnish bariatric surgery cohort (**Fig. 1a**). We generated longitudinal SAT snRNA-seq data from these 167 biopsies taken at four time-points, baseline (0m) (n=33), followed by a 4-6-week very low calorie diet (VLCD); operation (op) (n=68); 6 months (6m) (n=33) and 12 months (12m) (n=33) after the operation, resulting in a total of 137,061 quality control passing nuclei (see Methods). **Supplementary Data 1** shows the clinical characteristics of the 33 individuals with snRNA-seq data at all four time-points.

**Fig. 1:**
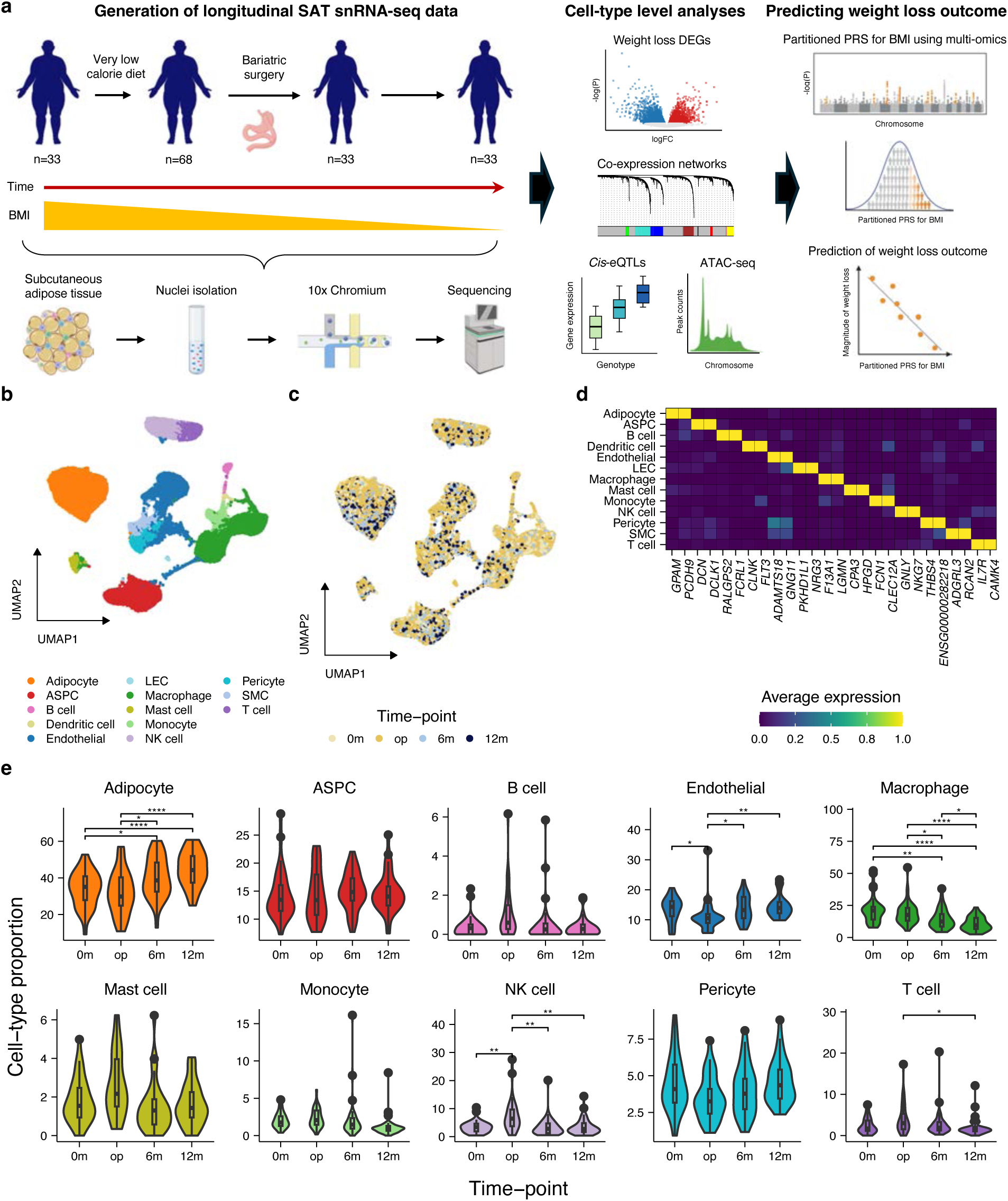
Longitudinal SAT snRNA-seq data from four time-points in the RYSA cohort reveals cell-type level differences by weight loss. **a** Schematic overview of the study design that uses longitudinal SAT snRNA-seq and adipocyte ATAC-seq data to construct multi-omics annotation-based partitioned PRS for BMI to predict weight loss outcome after bariatric surgery. **b,c** Uniform Manifold Approximation and Projection (UMAP) visualization of the 137,061 nuclei collected from 167 subcutaneous adipose tissue (SAT) biopsies across four time-points (0m, baseline; op, operation; 6m, 6 months after operation; and 12m, 12 months after operation) colored by the assigned cell-type (**b**) and originating biopsy time-point (**c**). **d** Average normalized expression of top protein coding unique cell-type marker genes used to annotate each population of cells. **e** Violin plots showing the proportion (%) of the 10 most frequent cell-types in the 33 individuals that had SAT biopsies collected from all four time-points. Center lines indicate the median cell-type proportions and boxes range from the 25^th^ to the 75^th^ percentiles. Cell-types with significant differences in proportions between the pairwise combinations of time-points are shown by Wilcoxon rank sum test. Asterisks indicate a significant *p*-value after Bonferroni adjustment (*, *p*.adj<0.05; **, *p*.adj<0.01; ***, *p*.adj<0.001). Exact *p*-values are available in Supplementary Data 3. ASPC indicates adipose stem and progenitor cells; ATAC-seq, assay for transposase-accessible chromatin sequencing; BMI, body mass index; DEGs, differentially expressed genes; LEC, lymphatic endothelial cells; NK, natural killer cells; PRS, polygenic risk scores; SMC, smooth muscle cells; and snRNA-seq, single nucleus RNA-sequencing. Fig. 1a created in BioRender. Lee, S. (2025) https://BioRender.com/ypln8cx

To establish single nucleus SAT transcriptomes of weight loss, we first clustered all 137,061 nuclei and used their unique cell-type marker genes (i.e., genes highly expressed in only one cell-type over the others) together with known SAT cell-type marker genes^8^ to classify them into 13 main cell-types under 5 broad cell-types, including adipocytes, adipose stem and progenitor cells (ASPCs), lymphoid cells, myeloid cells, and vascular cells (**Fig. 1b–d, Supplementary Fig. 1, and Supplementary Data 2**). We observed no clusters driven by sequencing batches or individual samples (**Supplementary Fig. 1a**). Instead, we detected substructures within the main cell-types by the time-point (**Fig. 1c, Supplementary Fig. 1e,f**), suggesting that weight loss is driving cell-type and subcell-type level transcriptome profiles.

### Cell-type composition changes by weight loss

We searched for differential cell-type composition by the weight loss both from the VLCD and bariatric surgery. We first observed significant differences in the mean cell-type proportions across the time-points for adipocytes, endothelial cells, macrophages, natural killer (NK) cells, and T cells using Friedman test (**Fig. 1e**). Among these cell-types, we further detected significant cell-type proportion differences between some time-points using Wilcoxon rank sum test (**Fig. 1e and Supplementary Data 3**). For example, adipocytes showed higher proportions in the two follow-up time-points when compared to the operation time-point (**Fig. 1e and Supplementary Data 3**). In contrast, immune cell-types, including macrophage, NK cells, and T cells, showed lower cell-type proportions in the follow-up time-points (**Fig. 1e and Supplementary Data 3**), suggesting attenuation of the obesity-induced low-grade inflammation following the drastic weight loss by bariatric surgery. No significant changes in the adipocyte, macrophage, and T cell proportions were observed after the weight loss by the preoperative VLCD alone.

### VLCD and surgery induced weight loss exhibit distinct changes in gene expression

We next investigated changes in cell-type level gene expression after weight loss by performing a pairwise differential expression (DE) analysis between time-points followed by functional enrichment analysis. In the DE analysis between the time-points op and 12m, where the largest mean delta BMI (−9.28 kg/m^2^ (SD=3.30)) was measured, the highest number of significant (FDR<0.05) differentially expressed genes (DEGs) were observed in adipocytes (n=4,003 genes), followed by ASPCs (n=1,090) and macrophages (n=691) (**Fig 2a and Supplementary Data 4**). In the gene ontology (GO) enrichment analysis, the adipocyte DEGs upregulated in op versus 12m, such as *ERRFI1*, *TGFBR2*, and *PTPRZ1*, were enriched for pro-inflammatory responses to obesity, including regulation of epithelial cell proliferation, angiogenesis, and cytokine-mediated signaling (**Supplementary Data 5**). In contrast, the adipocyte genes upregulated in 12m versus op, such as *ABCD1*, *ETFA*, *SESN2*, were enriched for lipid metabolism pathways, including fatty acid beta-oxidation, fatty acid homeostasis, and regulation of lipid catabolic process (**Fig. 2b and Supplementary Data 5**), in line with the use of SAT fat as fuel after the surgery. In ASPCs, DEGs upregulated at op versus 12m showed enrichment for potassium and sodium ion transport and bone morphogenetic protein (BMP), involved in signaling and response process for regulation of cell differentiation and apoptosis^10^ (**Fig. 2b and Supplementary Data 5**). Lastly, T cell genes upregulated in op versus 12m showed enrichment for T cell differentiation and activation, which supports the higher T cell proportions observed at op compared to 12m (**Fig. 2b and Supplementary Data 5**).

**Fig. 2:**
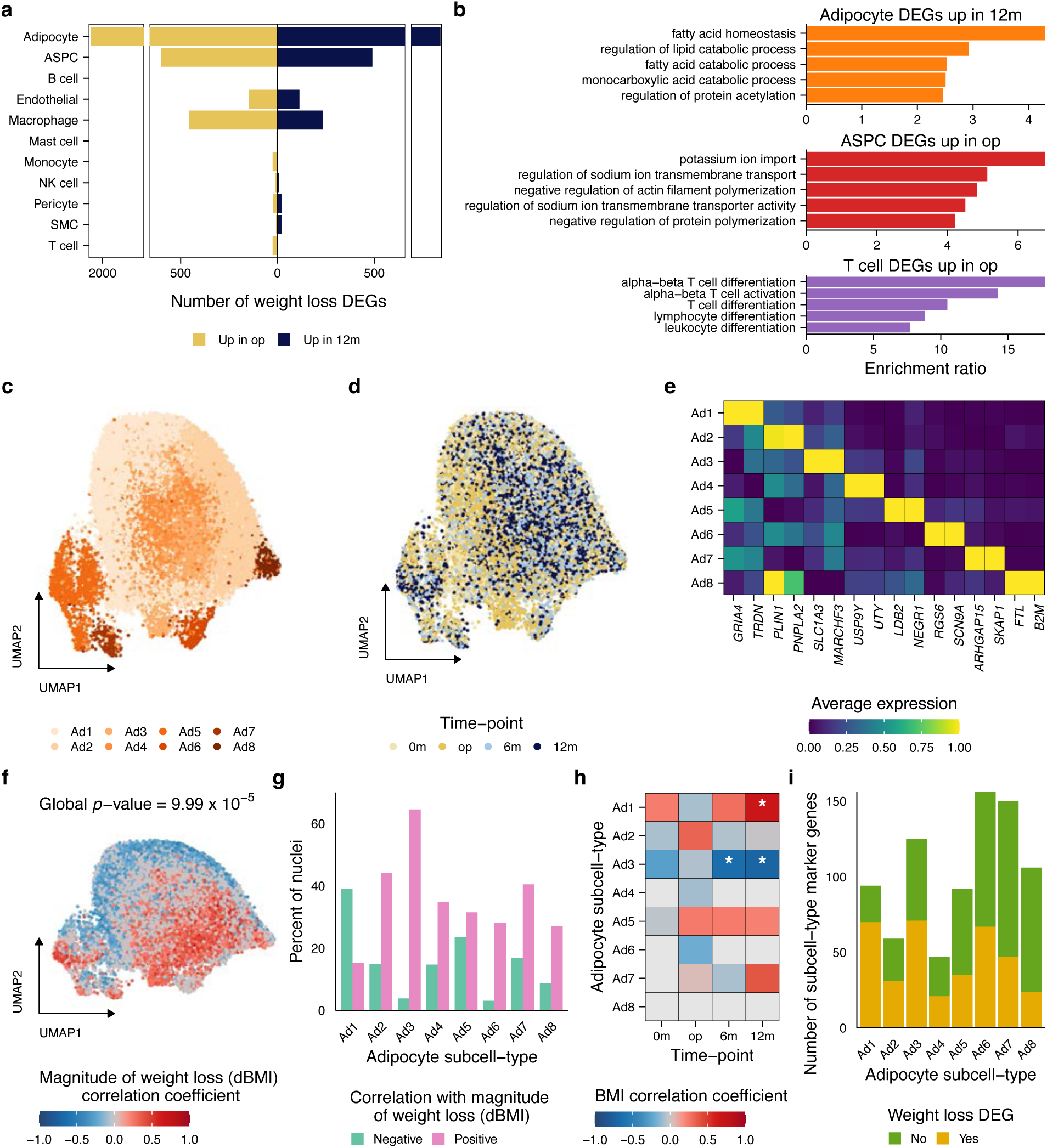
Adipocyte weight loss DEGs and subcell-types exhibit large heterogeneity. **a** Number of differentially expressed genes (FDR<0.05) between the op and 12m time-points per cell-type are shown colored by the time-point in which the genes are upregulated. **b** Top pathway enrichment results (FDR<0.05) for the genes differentially expressed between the op and 12m time-points. The full pathway enrichment results for all cell-type level DEGs are available in Supplementary Data 5. **c,d** Uniform Manifold Approximation and Projection (UMAP) visualization of adipocytes collected from the 33 individuals with SAT snRNA-seq data available at four time-points (0m, baseline; op, operation; 6m, 6 months after operation; and 12m, 12 months after operation) (n=132 SAT biopsies), colored by subclusters (**c**) and the originating biopsy time-point (**d**). **e** Average normalized expression of the top protein coding adipocyte subcell-type marker genes used to annotate each adipocyte subcell-type. **f** UMAP visualization of adipocytes colored by the correlation between adipocyte abundance and individuals’ magnitude of weight loss (delta BMI) using CNA (FDR<0.05). Adipocytes assigned to CNA neighborhoods with non-significant correlations (FDR≥0.05) are shown in gray. An adipocyte with a positive correlation indicates that it is assigned to a CNA neighborhood that has more nuclei from individuals with higher dBMI. **g** Percent of nuclei assigned to a CNA neighborhood that has significant correlation (FDR<0.05) with dBMI for each adipocyte subcell-type. **h** Heatmap showing correlations between adipocyte subcell-type proportions and body mass index (BMI) at four time-points (0m, baseline; op, operation; 6m, 6 months after operation; and 12m, 12 months after operation). Adipocyte subcell-types present in ≤20 individuals at each time-point were excluded from the analysis and are represented in gray. Asterisks indicate the *p*-value for Spearman correlation (*, Bonferroni adjusted *p*-value<0.05). **i** Number of subcell-type marker genes identified for each adipocyte subcell-type colored by whether they are weight loss DEGs. Ad indicates adipocytes; ASPC, adipose stem and progenitor cells; dBMI, delta body mass index; DEGs, differentially expressed genes; and FDR, false discovery rate.

After the VLCD induced weight loss (mean delta BMI of −1.73 kg/m^2^ (SD=1.21); n=33), we similarly observed that the greatest number of DEGs were obtained in adipocytes (n=3,725), followed by ASPCs (n=973) and macrophages (n=617) (**Supplementary Fig. 2a and Supplementary Data 4**). Interestingly, in the GO enrichment analyses, the adipocyte DEGs upregulated after VLCD were enriched for important adipocyte functions (**Supplementary Fig. 2b,c and Supplementary Data 5**), suggesting that adipocytes may be responding to VLCD by improving their energy homeostasis and increasing insulin sensitivity (**Supplementary Fig. 2b,c and Supplementary Data 5**). Overall, however, only 88 genes in adipocytes were DE in the same direction by both VLCD (0m vs op) and surgery (op vs 12m) induced weight loss while no other cell-type exhibited common DEGs in the same direction between the two modes of weight loss (**Supplementary Data 4**). These results likely reflect the fact that a much smaller mean weight loss was achieved by the VLCD when compared to the mean weight loss achieved by the bariatric surgery. Additionally, we observed just two adipocyte, two ASPC, and eight macrophage DEGs between the two post-surgery time-points (6m vs 12m) (**Supplementary Data 4**). Thus, to highlight the effects of the largest weight loss on SAT transcription profiles, we focus on the analyses between time-points op and 12m (mean delta BMI of −9.28 kg/m^2^ (SD=3.30)) in the remaining parts of the manuscript.

### Adipocyte heterogeneity drives subcell-type composition

The high resolution of our data revealed a wide-spread subcellular heterogeneity by weight loss, especially in adipocytes, which also exhibited the highest number of weight loss DEGs (**Supplementary Fig. 1)**. To investigate how the changes in the adipocyte transcription profiles by weight loss are reflected by variation in subcellular composition, we performed clustering in adipocytes and identified eight adipocyte subcell-types, each with distinct unique subcell-type marker genes (**Fig. 2c–e and Supplementary Data 6**). The GO analysis on these unique marker genes of each subcell-type revealed distinct terms significantly enriched between the adipocyte subcell-types (**Supplementary Fig. 3a and Supplementary Data 7**). For example, the subcell-type Ad2 showed enrichment for regulation of triglyceride metabolic process, with higher expression of *PNPLA2*, *SLC27A1*, *SREBF1*, and *DGAT2*, whereas the subcell-type Ad3 was predominantly enriched for regulation of muscle hypertrophy, muscle cell differentiation, and regulation of lipid localization and transport (**Supplementary Fig. 3a and Supplementary Data 7**).

We next compared our subcell-types by weight loss to those identified in the white adipose single cell tissue atlas without weight loss by Emont et al.^8^ (**Supplementary Fig. 3b**). Although the comparison showed some overlap in adipocyte subcell-types between the two cohorts, we observed overall distinct subcell-type populations, likely explained by the presence of visceral adipose tissue (VAT) in the previous atlas and the heterogeneity of subcell-types reflecting the phenotypic differences across the donors between the two cohorts, such as age, sex, and ethnicity, as well as the changes in BMI (dBMI) given that the longitudinal SAT nuclei in this study were sampled from individuals with extreme obesity before and after their weight loss (**Supplementary Fig. 3c–f**). In contrast, we observed overall concordance in the functionally distinct ASPC, lymphoid, myeloid, and vascular subcell-types (**Supplementary Fig. 4 and Supplementary Data 6,7**), suggesting that adipocyte subtypes react most profoundly to weight loss.

To further investigate whether the adipocyte subcell-types are associated with the degree of weight loss (i.e., dBMI) between the op and 12m time-points, we applied the co-varying neighborhood analysis (CNA) tool^11^. We observed an overall significant heterogeneity in adipocytes by dBMI (**Fig. 2f**). Among the adipocytes assigned to the neighborhoods by CNA that were significantly associated with dBMI, Ad3 had the greatest percentage of its nuclei (3,683 out of 3,900 nuclei) with positive correlation, indicating an increase in Ad3 abundance for individuals with greater magnitude of weight loss (**Fig. 2g**). In contrast, Ad1 had the greatest percentage of its nuclei (5,075 out of 7,061) with negative correlation, indicating a decrease in Ad1 abundance for individuals with greater magnitude of weight loss (**Fig. 2g**). Furthermore, the proportion of the adipocyte subcell-type Ad3 was significantly negatively correlated with BMI at the two post-surgery time-points, 6ms (Rho=−0.648; *p*.adj=8.56×10^−4^) and 12m (Rho=−0.707; *p*.adj=1.16×10^−4^) and that of Ad1 was significantly positively correlated with BMI at the 12m time-point (Rho=0.662; *p*.adj=9.49×10^−4^), supporting the observed differences in these subcell-type abundance based on individuals’ weight after bariatric surgery (**Fig. 2h**). Interestingly, the subcell-types Ad1 and Ad3 also exhibited the highest number weight loss DEGs among their unique marker genes (**Fig. 2i**). Taken together, these results indicate that the distinct obesity and weight loss related transcriptome profiles likely drive the subclustering in adipocytes. Thus, we focused on adipocytes in the remaining parts of the manuscript to further investigate their relationship with individuals’ weight loss outcome.

### Adipocyte weight loss DEGs form co-expression networks

To assess whether the identified adipocyte subcell-types reflect underlying cell-type level networks, we constructed cell-type level weighted gene co-expression networks at op and projected them to the 12m time-point data using hdWGCNA^12^ (**Supplementary Data 8**). We identified nine adipocyte co-expression modules (i.e., networks) with distinct functions, of which seven were enriched for unique marker genes of one to two adipocyte subcell-types (**Fig. 3a and Supplementary Data 9,10**). For example, Ad network III displayed several significant pathways related to the regulation of and response to cytokines, innate immune system, and inflammation, including genes such as *ANXA1*, *IL6ST*, and transcript factors *FOXO3* and *JUN*, all of which were upregulated at the operation time-point (**Fig. 3b and Supplementary Data 9**). This result suggests that Ad network III harbors a set of co-expressed genes in adipocytes that react to obesity by promoting innate immune response and inducing low-grade inflammation. In contrast, Ad network VIII, which was enriched for the unique marker genes of weight loss associated subcell-type Ad3, comprises 470 weight loss DEGs, including 8 genes upregulated at the 12m time-point, centered around sequestering of triglycerides and fatty acid oxidation, i.e., SAT fat burning, such as *ETFA*, *ECHDC2*, and *PRKAG2* (**Fig. 3c and Supplementary Data 9**). Overall, the network genes in Ad network VIII showed significant enrichment for pathways related to the key adipose tissue function of lipid storage and mobilization, which included these pathways of sequestering of triglyceride and fatty acid oxidation. In addition, module preservation analysis revealed that Ad network VIII is highly preserved at the 12m time-point (**Fig. 3d**). These results demonstrate that the adipocytes undergo coordinated upregulation of genes for enhanced adipocyte metabolism following the weight loss from bariatric surgery, thus reflecting changes towards metabolically healthy adipocytes. Notably, a transcription factor (TF) *ESR1* that targets *PRKAG2* was also found in the Ad network VIII, supporting the notion that TFs and their target genes show subcell-type level coordinated expression (**Supplementary Data 8**).

**Fig. 3:**
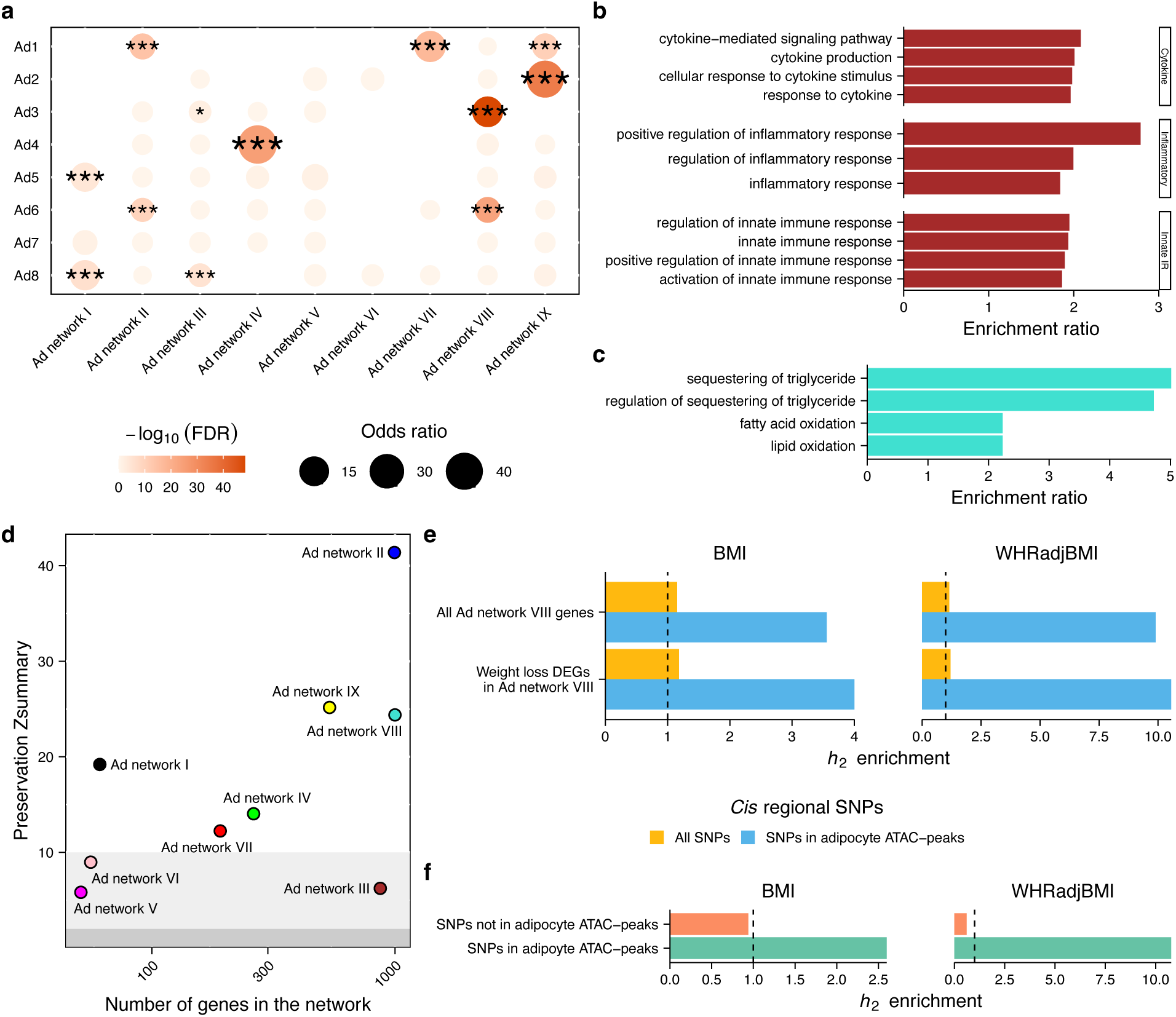
Expression of adipocyte weight loss genes is tightly coordinated in adipocyte networks. **a** Dot plot showing enrichment of unique adipocyte subcell-type marker genes for each adipocyte network. The dots are colored by significance of enrichment (−log_10_FDR) and their size represents an odds ratio of the overlap. Significantly enriched gene sets by Fisher’s exact test are highlighted by asterisks based on FDR (*, FDR<0.05; **, FDR<0.01; and ***, FDR <0.001). The full summary statistics from the Fisher’s exact tests are available in Supplementary Data 10. **b** Selected gene ontology (GO) biological process terms related to the regulation of and response to cytokines, inflammation, and innate immune response that were enriched (FDR<0.05) among the Ad network III genes. **c** Selected GO biological process terms related to sequestering of triglycerides and fatty acid oxidation that were enriched (FDR<0.05) among the Ad network VIII genes. The full pathway enrichment results for all network genes are available in Supplementary Data 9. **d** Adipocyte network preservation Z summary scores of the op time-point on the 12m time-point are shown plotted by the size of each network. The background of the scatterplot is shaded based on the preservation of the networks, where dark gray (Z≤2) indicates not preserved, light gray (2<Z≤10) moderately preserved, and no shade (Z>10) highly preserved, respectively. **e** Bar plots showing significant (*p*-value<0.05) enrichment of heritability, as measured by heritability ratio (*h_2_*), for BMI and WHRadjBMI by *cis* regional SNPs of all or weight loss DEGs in Ad network VIII. Blue color represents the SNPs landing in adipocyte open chromatin regions. Vertical dashed lines indicate the threshold of no enrichment or depletion of heritability in the tested genomic regions. **f** Bar plots showing significant (*p*-value<0.05) enrichment or depletion of heritability, as measured by heritability ratio (*h_2_*), for BMI and WHRadjBMI by SNPs landing in or not landing in adipocyte open chromatin regions. Vertical dashed lines indicate the threshold of no enrichment or depletion of heritability in the tested genomic regions. Ad indicates adipocytes; ATAC, assay for transposase-accessible chromatin sequencing; BMI, body mass index; FDR, false discovery rate; innate IR, innate immune response; SNP, single nucleotide polymorphism; and WHRadjBMI, waist-to-hip ratio adjusted for BMI.

To understand the relationship between the co-expression networks and genetic risk for BMI, we first used MAGMA^13^ and observed a significant enrichment for the BMI risk loci only with the genes in Ad network VIII (beta=0.106; *p*-value=9.90×10^−3^). Next, using linkage disequilibrium-score regression (LDSC)^14^ to investigate the relationship between Ad network VIII gene regions and heritability for BMI, we found that the *cis* regional SNPs of the Ad network VIII genes are significantly enriched for the heritability of BMI (**Fig. 3e and Supplementary Data 11**). When we further subset for the *cis* regional SNPs of Ad network VIII genes that are also weight loss DEGs, we also observed a significant heritability enrichment (**Fig. 3e and Supplementary Data 11**). The heritability of waist-to-hip ratio adjusted for BMI (WHRadjBMI), a well-established proxy of abdominal obesity^15^, was also enriched among the *cis* regional SNP sets of all genes and weight loss DEGs in Ad network VIII (**Fig. 3e and Supplementary Data 11**). Next, we overlapped the *cis* regional SNPs with adipocyte ATAC-peaks since SNPs landing in open chromatin regions are enriched for heritability of multiple CMD traits^16–18^. We observed that SNPs landing in adipocyte open chromatin regions were significantly enriched for the heritability of both BMI and WHRadjBMI whereas those landing outside the peaks were depleted for the heritability of both traits (**Fig. 3f**). Furthermore, for both all and weight loss DE gene sets in Ad network VIII, their *cis* regional SNPs landing in adipocyte ATAC-peaks showed stronger enrichment for heritability of BMI and WHRadjBMI (**Fig. 3e and Supplementary Data 11**). Noteworthy, this network was also highly preserved in independent SAT snRNA-seq data that we generated from 35 RYSA participants’ op time-point biopsies that were not included in our main analyses, thus ensuring independent replication of the network (**Supplementary Fig. 5**). Altogether, these results indicate that the adipocyte open chromatin regions around weight loss DEGs in Ad network VIII are enriched for genetic variants associated with obesity traits.

### Cell-type level time-point-shared and -specific *cis* regulation of adipocyte weight loss DEGs

To investigate which weight loss DEGs or adipocyte expressed genes are under genetic regulation, we performed a cell-type level *cis*-eQTL mapping of all expressed genes in adipocytes across the four time-points. We applied multivariate adaptive shrinkage with mashr (see Methods) to further identify time-point-shared and -specific eQTLs. In adipocytes, we identified 7,217 genes with *cis*-eQTL SNPs (i.e., eGenes) (local false sign rate<0.05) in at least one time-point, which included 2,222 (56% of the) weight loss DEGs (op vs 12m) (**Fig. 4a**).

**Fig. 4:**
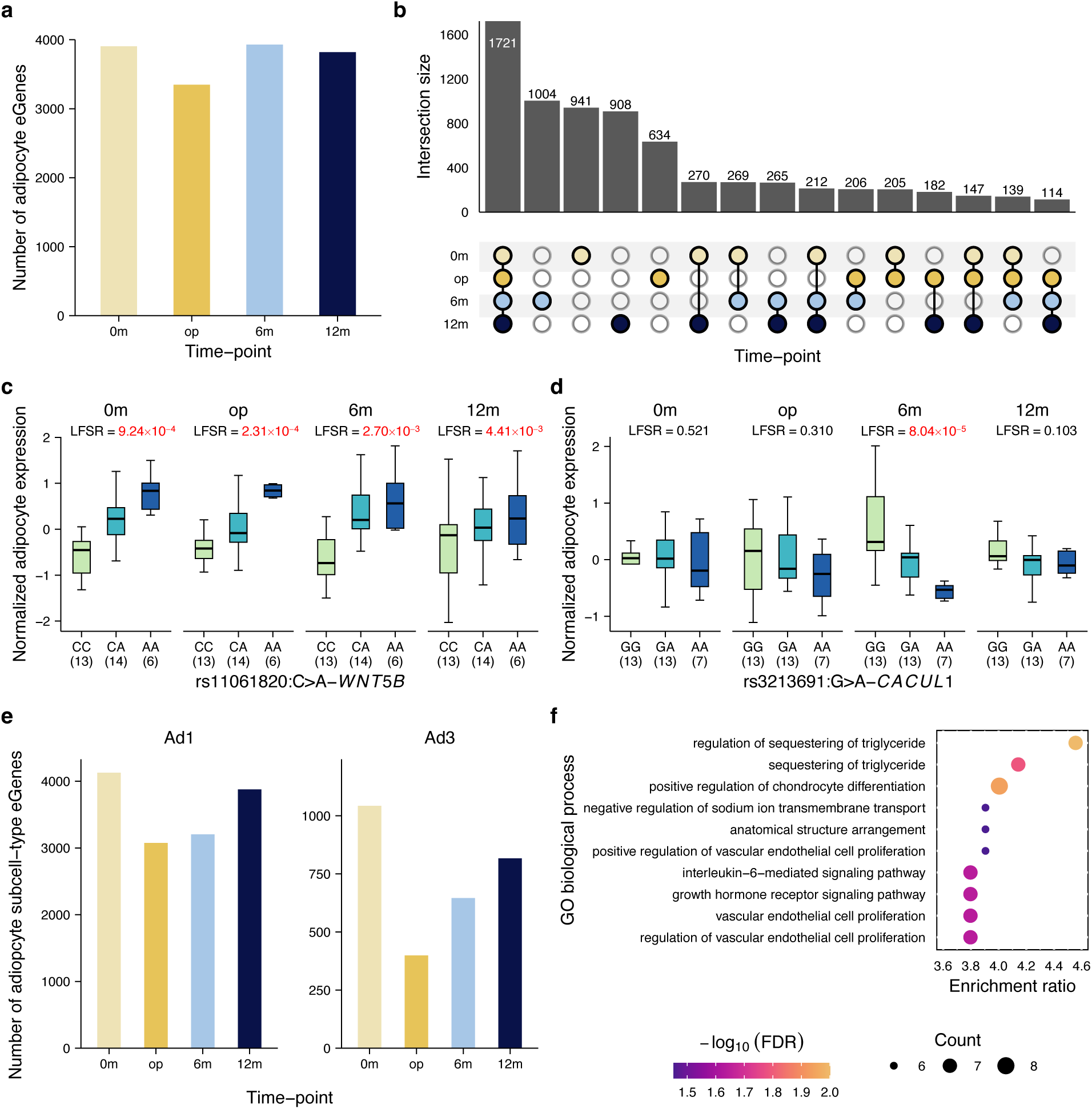
Cell-type and subcell-type level *cis*-eQTL analyses discover time-point level genetic regulation of adipocytes and adipocyte subcell-types. a Bar plots showing the numbers of genes with significant (LFSR<0.05) adipocyte *cis*-eQTL SNPs (i.e., eGenes) at each time-point (0m, baseline; op, operation; 6m, 6 months after operation; and 12m, 12 months after operation). **b** Upset plot showing the number of time-point-shared and -specific adipocyte eGenes for all possible combinations of time-points in adipocytes. The top bar plots represent the number of adipocyte eGenes and the bottom dots indicate the time-point combinations. **c,d** Examples of a time-point-shared adipocyte *cis*-eQTL SNP-gene pair, rs11061820:C>A-*WNT5B* (**c**), and of a 6m, time-point-specific adipocyte *cis*-eQTL SNP-gene pair, rs3213691:G>A-*CACUL1* (**d**). Box plots show normalized adipocyte expression by the genotype group, center lines indicate the median, and boxes range from the 25^th^ to the 75^th^ percentiles. **e** Bar plots showing the numbers of genes with significant (LFSR<0.05) subcell-type level *cis*-eQTL SNPs (i.e., eGenes) at each time-point for the adipocyte subcell-types, Ad1 and Ad3. **f** Dot plot showing enrichment of the top ten gene ontology (GO) biological process terms by the eGenes discovered in the adipocyte subcell-type A3, based on the significant (FDR<0.05) enrichment ratio. The dots are colored by significance of enrichment (−log_10_FDR) and their size represents the number of overlapping genes with each term. Ad indicates adipocyte subcell-type; FDR, false discovery rate; LFSR indicates the local false sign rate; and SNP, single nucleotide polymorphism.

Evaluating across the time-points, we detected 1,712 (23%) shared and 634-1,004 (9-14%) time-point specific eGenes (**Fig. 4b**). For example, the gene *WNT5B*, involved in preadipocyte to adipocyte differentiation^19^ (i.e., adipogenesis), showed consistent *cis*-eQTL effects across all four time-points (**Fig. 4c**), suggesting that its allele-specific role in adipogenesis is maintained throughout the weight loss. In contrast, the *cis*-eQTL variant rs3213691 showed a 6m time-point-specific effect on adipocyte expression of *CACUL1* (**Fig. 4d**). *CACUL1* encodes a protein that interacts and represses the transcriptional activity of PPARγ, a known master regulator of adipogenesis, and consequently *CACUL1* inhibits preadipocyte differentiation to adipocyte^20^. These two examples of time-point-shared vs -specific *cis*-eQTL effects highlight the heterogeneity in the genetic regulation of adipogenesis by adipocytes in response to weight loss. We assessed the robustness of the *cis*-eQTL results by evaluating the replication of the adipocyte *cis*-eQTL SNPs with the bulk SAT *cis*-eQTLs from the phenotypically similar Finnish bariatric surgery cohort, KOBS^21^, and the Genotype-Tissue Expression (GTEx) project^22^ (**Supplementary Data 12**). As the KOBS *cis*-eQTL analysis was done on the SAT bulk RNA-seq data from the bariatric surgery operation time-point, we focused on the replication of the adipocyte eGenes from the RYSA op time-point results. Focusing on the SNP-gene pairs that were tested in both RYSA and KOBS cohorts, we observed that 44.6% of the RYSA adipocyte eGenes shared at least one *cis*-eQTL SNP with the KOBS SAT bulk eGenes (q-value<0.05 in the same direction of effect). Additionally, the replication rate in the KOBS was 35.5% for the lead *cis*-eQTL SNPs of the RYSA adipocyte eGenes. Finally, we also calculated the ν_1_ statistic^23^ to estimate the proportion of the RYSA adipocyte lead SNP-gene pairs that replicated in the KOBS bulk and observed ν_1_=0.644, i.e., 64.4% replicated. In GTEx, 42.5% of the RYSA adipocyte eGenes shared at least one *cis*-eQTL SNP with the SAT bulk eGenes (q-value<0.05 in the same direction of effect), 25.3% of the lead *cis*-eQTL SNPs of the RYSA adipocyte eGenes replicated, and by calculating the ν_1_ statistic, we observed a ν_1_ of 0.608, i.e., 60.8% replicated. These replication rates are comparable to or better than the previously reported replication rates of cell-type level *cis*-eQTL results from solid tissues when comparing with bulk tissue *cis*-eQTL results^23–25^, suggesting that the difference in the eGenes discovered between the RYSA SAT adipocyte and KOBS and GTEx SAT bulk *cis*-eQTL analyses may be contributed by the difference in the single-cell vs bulk resolution of the data sets (i.e., differences in the modality).

### Adipocyte subcell-type eGenes largely overlap with adipocyte eGenes

To evaluate whether *cis* regulation at a subcell-type level captures the distinct biological functions of adipocyte subcell-types, we further performed adipocyte subcell-type level *cis*-eQTL mapping using mashr, limiting these analyses to the two weight loss and BMI-associated adipocyte subcell-types, Ad1 and Ad3 (**Fig. 4e**). We discovered 6,933 eGenes in at least one time-point for Ad1 (4,128 at 0-month, 3,074 at operation, 3,204 at 6-month, and 3,878 at 12-month, respectively) and 1,692 eGenes in at least one time-point for Ad3 (1,043 at 0-month, 399 at operation, 646 at 6-month, and 817 at 12-month, respectively). Of the 6,933 and 1,692 eGenes for Ad1 and Ad3, 4,588 (66.2%) and 1,248 (73.8%) overlapped with the 7,217 eGenes discovered in all adipocytes, indicating that many of the subcell-type eGenes were captured by the main adipocyte *cis*-eQTL analyses. Interestingly, we observed significant enrichment of functional pathways for the 1,692 eGenes from Ad3 (FDR<0.05), with the top enriched pathways being functionally important adipocyte pathways: sequestering of triglyceride and its regulation (**Fig 4f and Supplementary Data 13**). No pathway enrichments were observed with the Ad1 eGenes. Notably, the genes involved in these adipocyte pathways of Ad3 eGenes included *ABHD5*, *LPL*, *PNPLA2*, and *PPARA*, all of which are eGenes observed only in the subcell-type Ad3 but not in the full adipocytes. Of these four genes, *ABHD5* is a highly expressed unique subcell-type marker gene of Ad3, suggesting that the high expression of *ABHD5* uniquely in Ad3 contributes to the distinct functions of this subcell-type and likely explains the lack of *cis*-eQTL results for this gene in the full adipocyte data. In contrast, the other three genes were also expressed in other adipocyte subcell-types but their genetic regulation were only observed in Ad3 rather than in the full adipocyte data. Overall, these subcell-type *cis*-QTL results suggest that the genes underlying the adipocyte enriched functions of Ad3 are under genetic control and that subcell-type level *cis*-eQTL analyses can further refine genetic regulations of biologically distinct subcell-types.

### Network PRS for BMI is a significant predictor of weight loss outcome

As a large proportion of weight loss DEGs are under genetic regulation and *cis* regional SNPs are enriched for heritability of obesity traits, we hypothesized that individuals’ genetic risk for obesity may contribute to interindividual variation in the weight loss outcome achieved by bariatric surgery. To assess the relationship between the individuals’ degree of weight loss after bariatric surgery and their genetic risk for obesity, we first constructed genome-wide polygenic risk scores (PRS) for BMI a Finnish population cohort, METSIM^26^ (n=8,254 unrelated with mean BMI of 27.27 kg/m^2^; SD=4.15), and was subsequently jointly applied to the METSIM, RYSA (n=68), and an independent Finnish bariatric surgery cohort, KOBS (n=170)^27^ (see Methods). We first observed that the genome-wide BMI PRS is significantly higher in RYSA and KOBS than in METSIM (*p*.adj=5.4×10^−10^ and 2×10^−16^, respectively), while not different between the two bariatric surgery cohorts (*p*.adj=1) by the Wilcoxon test (**Fig. 5a**). We further found that the mean BMI PRS in the RYSA and KOBS cohorts are in the 77^th^ and 79^th^ percentile of the METSIM PRS percentiles, respectively (**Fig. 5a**). This implies that in line with a previous study^2^, these RYSA and KOBS individuals who underwent bariatric surgery are already at genetically higher risk for obesity as measured by BMI PRS even when compared to METSIM that has a mean BMI at the overweight level.

**Fig. 5:**
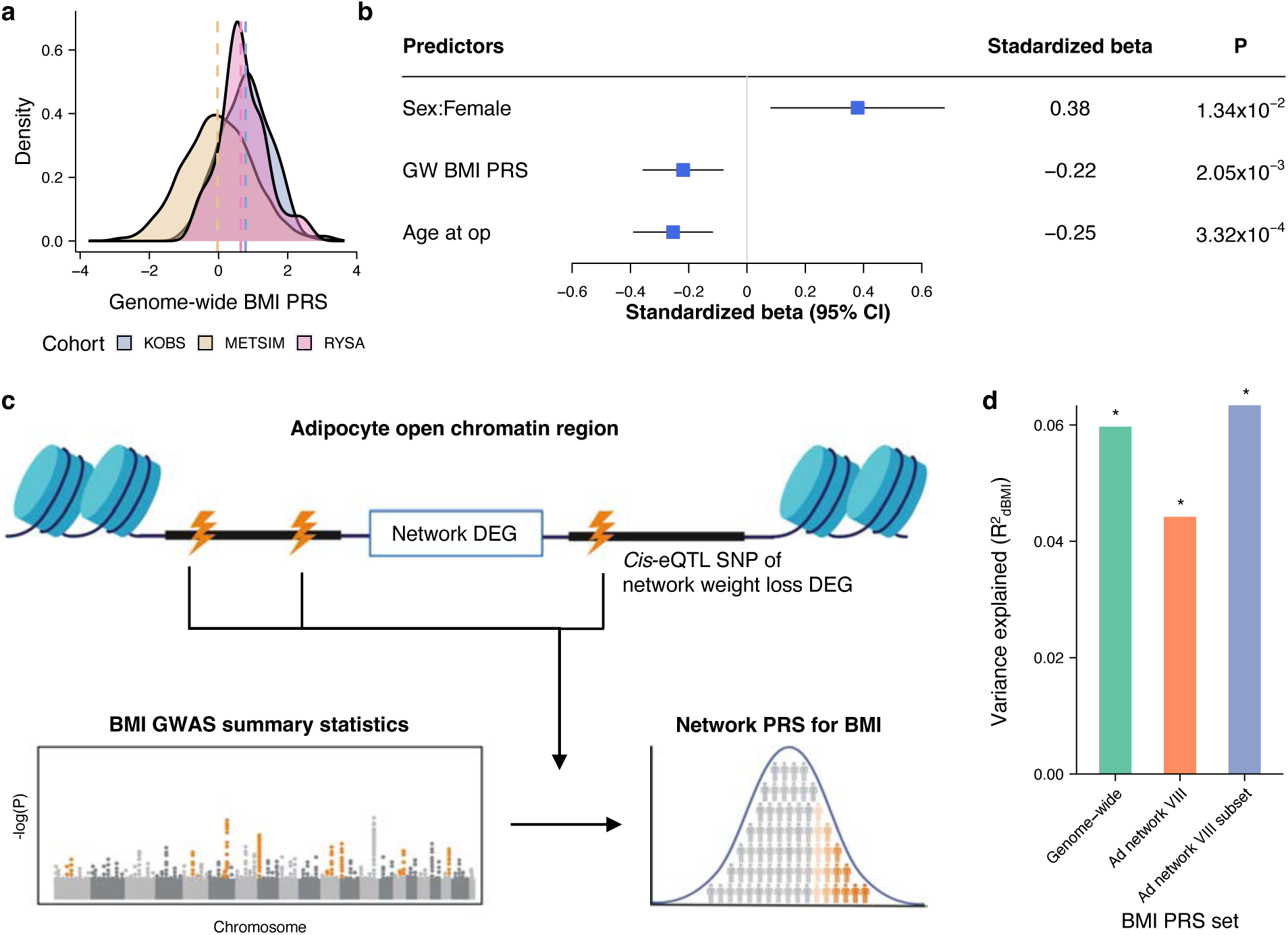
Network polygenic risk scores for obesity significantly predict weight loss outcome. **a** Distributions of genome-wide (GW) BMI polygenic risk scores (PRSs), colored by the METSIM, KOBS, and RYSA cohorts. **b** Forest plot showing the standardized effect of age, sex, and genome-wide BMI PRS on the weight loss outcome (dBMI), measured by the change in BMI from the time of operation to the 12-month follow-up, while controlling for the baseline BMI, in the KOBS bariatric surgery cohort (n=170). The positive standardized beta values indicate a greater magnitude of BMI change. **c** Schematic overview of the SNP selection from the adipocyte open chromatin regions of the network DEGs for the construction of network PRSs (see Methods). **d** Variance explained in the BMI change between the operation and 12-month follow-up time-points (dBMI) in the KOBS bariatric surgery cohort by the genome-wide and network PRS sets. Asterisks indicate a significant *p*-value for variance explained in dBMI by each PRS set (*, *p*-value<0.05). Ad indicates adipocytes; BMI, body mass index; DEGs, differentially expressed genes; eQTL; expression quantitative trait loci; GWAS, genome-wide association study; and SNP, single nucleotide polymorphism. Fig. 5c created in BioRender. Lee, S. (2025) https://BioRender.com/61j5nbt.

We next investigated whether the individuals’ age (at op), sex, and genome-wide BMI PRS explain interindividual variation in the weight loss outcome. Due to the small sample size of the RYSA cohort, we focused these analyses on an independent Finnish bariatric surgery cohort, KOBS (n=170). We applied a linear model to predict the BMI change (dBMI) between op and 12m using the genome-wide BMI PRS, sex, and age as the predictors, while including BMI at op and the first 10 genetic PCs as covariates. We discovered age and sex, as well as individuals’ genome-wide BMI PRS as significant predictors of weight loss, where the BMI PRS explains 5.97% of the variation (R^2^) in dBMI (**Fig. 5b**).

Next, we assessed whether partitioned network PRS built using weight loss DEGs in the functionally enriched network of the RYSA SAT snRNA-seq data also predict dBMI in KOBS, hypothesizing that the regional variants of these cell-type level DEGs reacting to weight loss could further refine the genetic component regulating the interindividual variability of weight loss. We focused on the *cis* regional SNPs of weight loss DEGs in Ad network VIII that land in adipocyte open chromatin regions as these were significantly enriched for heritability of obesity traits. We further subset for *cis*-eQTL SNPs of these weight loss DEGs as *cis*-eQTL SNPs are more likely to be disease associated and account for stronger enrichment of disease heritability^28–30^. Thus, we constructed partitioned network BMI PRS with the *cis*-eQTL SNPs of the weight loss DEGs in Ad network VIII that land in adipocyte open chromatin regions (**Fig. 5c**). We discovered that the network BMI PRS of Ad network VIII (n=147 SNPs) significantly predict dBMI (*p*-value=8.15×10^−3^) and are enriched for variance explained in dBMI, explaining 4.42% (*p*_perm10,000_=9.40×10^−3^) of the variation (R^2^) in dBMI (**Fig. 5d and Supplementary Data 14,15**). When we further subset the Ad network VIII genes to the DEGs upregulated in op (n=45 genes), the network BMI PRS explained 6.34% (*p*-value=1.44×10^−3^) of the variation in dBMI (*p*_perm10,000_=1.70×10^−3^) (**Fig. 5d and Supplementary Data 14,15**). Thus, this network PRS of Ad network VIII subset (n=45 genes) captures a similar amount of variation in dBMI as the genome-wide BMI PRS (R^2^=5.97%) (**Fig. 5d and Supplementary Data 15**), supporting the conclusion that it comprises important weight loss genes.

The Ad network VIII subset BMI PRS set included 45 adipocyte eGenes from Ad network VIII that were also DEGs upregulated at the op time-point. Of these genes, 12 genes (*ABCA6*, *CNNM2*, *COMMD10*, *CRADD*, *EYA4*, *JMJD1C*, *MITF*, *PRR5L*, *RBM47*, *RBMS1*, *SLC39A11*, and *SNRK*) had strong to compelling genetic support for their involvement in anthropometric traits, such as BMI and waist-to-hip ratio as well as serum triglycerides, based on the Human Genetic Evidence (HuGE) scores, calculated using previous human genetic association studies^31^. Taken together, our longitudinal SAT snRNA-seq data from the RYSA cohort identified a set of co-expressed weight loss genes containing *cis* regulatory variants that significantly contribute to interindividual variability in weight loss after bariatric surgery and provide potential therapeutic targets for treatment of obesity.

## Discussion

Despite the fast-growing number of individuals with obesity worldwide, cell-type level responses to a large weight loss in subcutaneous adipose tissue (SAT), the main body tissue expanding and buffering against obesity, have remained elusive. This is mostly due to small sample sizes of existing single cell data and the lack of longitudinal omics cohorts. Consequently, little is known about genes underlying interindividual variability in weight loss at the single cell resolution.

Here we present a comprehensive longitudinal single nucleus transcriptomic profiling of SAT during weight loss from a Finnish bariatric surgery cohort.

Our temporal study of weight loss discovered that the SAT main and subcell-types undergo profound changes in their abundance and gene expression in response to weight loss, demonstrating physiological changes in SAT, induced by bariatric surgery. We then utilized the weight loss responding DEGs to first discover sets of co-expressed DEGs through a cellular co-expression network analysis in each cell-type. Next, we discovered genetic regulators, i.e., *cis*-eQTL variants, of the weight loss responding network DEGs that are either shared across the time-points or specific to a time-point. By integrating with adipocyte ATAC-seq data in the DEG *cis* regions, we constructed regional BMI polygenic risk scores (PRSs) using the *cis* regional variants of the weight loss responsive network DEGs. These PRSs both predict the weight loss outcome of bariatric surgery and are enriched for variance in the weight loss when compared to 10,000 permutations of randomly selected, size matched SNP sets from the genome. Taken together, using longitudinal SAT snRNA-seq data, we identify specific *cis* regulatory variants and their target genes, which respond to weight loss and predict the interindividual variability in the magnitude of weight loss.

Transcriptional changes in SAT by weight loss have been demonstrated before^7,32,33^; however, these studies were limited to bulk tissue data, masking the wide-spread cell-type heterogeneity present in SAT^8,34^ that is known to confound bulk RNA-seq results^35^. Our study demonstrates both the cross sectional (i.e., time-point level) and longitudinal cellular heterogeneity of SAT and discovers cell-type level DEGs by weight loss, thus providing insight into the cellular and molecular mechanisms underlying the responses of SAT to a large change in weight at the cell-type resolution. Our data revealed a substantial number of cell-type level DEGs by weight loss after the bariatric surgery, of which most are observed in adipocytes, the key SAT function-related cell-type. Our cell-type level co-expression network and pathway enrichment analyses found that even the adipocyte DEGs upregulated at the operation time-point are largely involved in immunological and apoptosis pathways rather than normal adipocyte functions, reflecting the low-grade inflammation ongoing in obese SAT. However, notably, healthy SAT function-related pathways appear among the adipocyte DEGs upregulated at the 6m and 12m time-points after weight loss.

Adipocytes in SAT have been shown to exhibit heterogeneity based on their cellular phenotypic characteristics^36–39^. A recent adipose meta single cell study^39^ also showed that adipocyte snRNA-seq data poorly integrate across studies and exhibit limited adipocyte marker gene reproducibility with the adipocytes from the previous adipose single cell atlas^8^. Similarly, we observed differences in the adipocyte subcell-types compared to the subcell-types from the previous adipose single cell atlas^8^. In our data, we found a significant heterogeneity in adipocytes based on individuals’ weight loss outcome by bariatric surgery as well as adipocyte subcell-types reflecting differences in individuals’ BMI. Thus, the adipocyte subcell-types from our longitudinal weight loss study support the notion that adipocyte heterogeneity reflects the underlying differences in the donors’ phenotypes.

We focused on the genes that significantly changed in expression between the op and 12m time-points to capture the maximum weight loss effect between these two time-points. Their set of DEGs showed very small overlap with the DEGs observed by VLCD between the 0m and op time-points, where only an average of −1.73 kg/m^2^ (SD=1.21) change in BMI was observed compared to an average of −9.28 kg/m^2^ (SD=3.30) change in BMI between the op and 12m time-points. It is possible that not all individuals fully complied with the strict VLCD diet, and biological heterogeneity may also have a large impact on this result. Thus, the difference between the 0m-op and op-12m DEGs might be reflecting the magnitude of weight loss achieved using the two types of obesity management. Furthermore, individuals still exhibit class III obesity after the VLCD with the mean BMI of 43.53 kg/m^2^ (SD=5.93), which suggests that SAT may continue to react to the obese environment when compared to SAT after the large weight loss by bariatric surgery. In addition, bariatric surgery induces changes in the gut microbiota composition^40^, which is known to be associated with adipose gene expression^41^. This suggests that alterations of the gut microbiome induced by diet alone versus bariatric surgery may differ, which might also contribute to the observed specific DEGs.

Our data extends beyond the previous atlas^8^ of SAT not only by characterizing the longitudinal single nucleus transcriptional landscape of SAT during weight loss but also by identifying regulatory variants associated with cell-type level gene expression and further classifying these variants by their context-specificity over the time-points. Due to small sample sizes of single cell SAT data, previous *cis*-eQTL analyses in SAT have so far been limited to bulk RNA-seq^42,43^. The high resolution and sufficient sample size of our data enabled the cell-type level *cis*-eQTL analyses, which identified over 7,000 genes under *cis* regulation. As genetic regulatory activities have been shown to be context-specific^44,45^, we further identified time-point-shared and -specific *cis*-eQTLs for the four main SAT cell-types. We found that the greatest number of eGenes were shared across the four time-points in adipocytes, supporting the notion that adipocytes appear to retain the genetic regulation of their key functions across the time-points. In summary, we discovered abundant cell-type level context-specific *cis* regulatory variants acting on the weight loss responsive eGenes.

Mapping *cis*-eQTLs at a subcell-type level can provide additional insight into genetic regulations that are specific to subpopulation of cells^24,46^; however, detecting eGenes in rare subcell-types is challenging as the number of eGenes discovered has been shown to correlate with the number of cells^24,46^. Consequently, we limited our subcell-type level *cis*-eQTL analyses to two weight loss and BMI-associated adipocyte subcell-types Ad1 and Ad3, the unique marker genes of which were enriched among the network genes in the functionally interesting Ad network VIII. We found that a large number of subcell-type eGenes overlapped with the full adipocyte eGenes, while also discovering subcell-type-specific eGenes. Noteworthy, the eGenes of Ad3 showed functional pathway enrichments, including the sequestering of triglycerides and its regulation, unlike the eGenes of the full adipocytes or Ad1. One of the genes involved in these pathways is Abhydrolase Domain Containing 5, Lysophosphatidic Acid Acyltransferase (*ABHD5*), that is an eGene only in Ad3 but not in Ad1 or the full adipocytes. *ABHD5* is a regulator of adipose triglyceride lipase-mediated lipolysis and a known cause of a rare monogenetic disorder (Chanarin-Dorfman syndrome), characterized by accumulation of triacylglycerol droplets in tissues^47,48^, further emphasizing the importance of this gene in lipid droplet formation, sequestering of triglycerides and its regulation in adipocytes. Overall, our subcell-type level *cis*-eQTL analysis reveals that the genes with biological functions in adipocyte subtypes are under distinct genetic control and presents the potential additional benefit of investigating SAT subcell-types for *cis*-eQTLs in the future studies with larger sample sizes.

Obesity is well known to be highly heritable yet polygenic^2,49^, and individuals with extreme obesity have previously been shown to exhibit higher PRSs for BMI^2^. Recent studies have used BMI PRSs constructed from relatively small subsets of previously known BMI GWAS variants to predict the weight loss outcome of individuals after bariatric surgery^50–52^. However, these studies showed conflicting results, possibly due to a mixed pool of SNPs used to construct the PRSs with no longitudinal functional genomics data to support the gene regions of interest for weight loss. Furthermore, the SNPs were limited to nearby genes of known BMI GWAS genes, suggesting that the genome-wide significant variants in the current cross-sectional BMI GWASs inadequately capture the genetic component of the weight loss outcome and in general warrant future large longitudinal GWASs on response outcomes. Here we utilized our longitudinal snRNA-seq data to first identify genes differentially expressed by weight loss and then performed co-expression network analysis to further classify the weight loss DEGs into different co-expression networks enriched in specific functional pathways. By then focusing on the BMI heritability enriched *cis*-eQTL SNPs that regulate the co-expressed weight loss DEGs and land in adipocyte open chromatin regions, we discover that their partitioned network PRSs for BMI significantly predict the BMI change in an independent Finnish bariatric surgery cohort, KOBS. Notably, one of these network PRSs explained variance in BMI change comparable to that by the genome-wide BMI PRS, emphasizing its importance as an individualized, functionally based genetic predictor of weight loss.

There are three TFs among the 45 DEGs in the BMI PRS network, that explained the most variance in the BMI change, including *KLF9*, a known regulator of mid-adipogenesis and *PPARG*^53^, which is in line with the key adipocyte functions significantly enriched in the entire Ad network VIII. Other notable genes in this PRS network are *JMJD1C* and *RBMS1*. A depletion of *Jmjd1c* has been shown to impair adipogenesis in 3T2-L1 murine preadipocytes^54^ and to be involved in promoting lipogenesis in liver^55^. Similarly, a previous mouse study observed high expression of *Rbms1* during early phase of 3T3-L1 murine preadipocyte differentiation (i.e., adipogenesis) as well as in mice on high-fat diet^56^. Additionally, they discovered that *Rbms1* is involved in altered cellular processes in adipocytes, including lipid metabolism^56^. These previous findings suggest that the regulation of adipogenesis and adipocyte metabolism may, at least in part, contribute to individuals’ weight loss outcome after bariatric surgery. Thus, our findings provide specific target genes with regulatory variants that can be used to better predict who might benefit more or less from bariatric surgery. Taken together, our network PRS results indicate the important role of genetics and specific genes along with an individual’s age and sex affecting the magnitude of change in BMI achieved after bariatric surgery.

Although we provide comprehensive single cell transcription profiles of SAT during weight loss, our study lacks visceral adipose tissue (VAT), shown to be linked to increased metabolic syndrome, insulin resistance, and cardiovascular risk^57^. However, obtaining longitudinal VAT data is very challenging in humans due to the invasive nature of VAT biopsies. In comparison, SAT biopsies are substantially less invasive, and the value of SAT data derives from the key role of SAT in its capacity to expand, thus providing a critical adaptive buffering mechanism against obesity^58^, making it a highly important tissue to study. In addition, we recognize that our study lacks other single cell level modalities, such as snATAC-seq, due to the existing technical challenges in generating large numbers of high-quality single cell level ATAC-seq data in frozen SAT biopsies. To partly circumvent this challenge, we utilized deep bulk ATAC-seq data, generated from differentiated human primary adipocytes, the major cell-type of SAT, which successfully identified regulatory *cis*-eQTL SNPs, landing in adipocyte open chromatin, that are significantly enriched for variance explained in the change of BMI during weight loss. Finally, we also recognize the limitation of our network PRS analyses in terms of the relatively small sample sizes of the two bariatric surgery cohorts. However, we believe that our unique longitudinal SAT snRNA-seq data set and our findings on transcriptional changes by weight loss, especially the underlying genetic components contributing to individuals’ weight loss, discovered through the network BMI PRS analyses, will provide a valuable resource to the scientific community to be included in future meta-analyses and for functionally targeted follow-up studies of weight loss.

## Methods

### Study cohorts

#### Roux-en-Y versus one-anastomosis gastric bypass (RYSA) cohort

In this study, we used 167 subcutaneous adipose tissue (SAT) biopsies, along with phenotype and genotype data from the longitudinal bariatric surgery cohort, Roux-en-Y versus one-anastomosis gastric bypass (RYSA)^9^. In RYSA, Finnish individuals with grade 3 obesity (n=130) were originally recruited in the Helsinki University Hospital, Helsinki, Finland for a randomized trial to receive either Roux-en-Y gastric bypass (RYGB) or One-anastomosis gastric bypass (OAGB) bariatric surgery, as described previously^9^. Clinical and laboratory examinations in RYSA were performed at the baseline, (0m), followed by a 4-6-week of very low calorie diet (VLCD) until the operation (op), and clinical and laboratory examinations were also performed during the 6-month (6m) and 12-month (12m) follow-ups (for clinical characteristics, see **Supplementary Data 1**), as described previously^9^. In this study, we generated single nucleus RNA-sequencing (snRNA-seq) data from 167 frozen SAT biopsies from the RYSA participants, resulting in 68 snRNA-seq samples at op as well as longitudinal snRNA-seq data from 33 of the 68 participants across all four time-points (0m, op, 6m, and 12m). All individuals were also genotyped using the Infinium Global Screening Array-24 v1 (Illumina). The RYSA study was approved by the Helsinki University Hospital Ethics Committee, and all participants provided a written informed consent. All research conformed to the principles of the Declaration of Helsinki.

#### Kuopio Obesity Surgery Study (KOBS) cohort

In this study, we used existing genotype and phenotype data from the longitudinal Kuopio Obesity Surgery Study (KOBS) cohort^27,42^. For KOBS, Finnish individuals (n=509) with obesity (BMI of ≥40 kg/m^2^ or 35 kg/m^2^ with a significant comorbidity) undergoing bariatric surgery were recruited in the University of Eastern Finland and Kuopio University Hospital, Kuopio, Finland, as described previously^27,42^. The KOBS study was approved by the Ethics Committee of the Northern Savo Hospital District, in accordance with the Declaration of Helsinki, and all participants provided written informed consent.

#### METabolic Syndrome In Men (METSIM) cohort

We used existing genotype and phenotype data from 8,257 unrelated Finnish males from the METabolic Syndrome In Men (METSIM) study, originally recruited at the University of Eastern Finland and Kuopio University Hospital, Kuopio, Finland^26,59^. The METSIM study was approved by the local ethics committee, in accordance with the Declaration of Helsinki, and all participants provided written informed consent.

#### UK Biobank cohort

We used phenotype and imputed genotype data from the UK Biobank (UKB) cohort. Detailed phenotype measurements were collected at time of recruitment for the full cohort, and a subset of the cohort were profiled for imaging data, including abdominal magnetic resonance imaging (MRI), in follow-up assessments^60–62^. Genotype data were obtained with either the Affymetrix or Applied Biosystems UK Biobank Axiom technology and imputed against the Haplotype Reference Consortium and the merged UK10K and 1000 Genomes phase 3 reference panel^61,62^.

Only the imputed genotype data from unrelated individuals of European ancestry with phenotype data available were included in our analyses. Data from UKB were accessed under application 33934.

#### Genotype quality control and imputation

We genotyped DNAs from RYSA and KOBS participants using the Infinium Global Screening Array-24 v1 (Illumina). The genotype data from each cohort were separately quality controlled using PLINK v1.9^63^ by excluding individuals with missingness >2%, unmapped and strand ambiguous SNPs, monomorphic SNPs, and variants with missingness >2% and Hardy-Weinberg Equilibrium (HWE) *p*-value<10^-6^. We also cross-checked reported sex of each individual with inferred biological sex imputed using the ‘--sex-check’ function in PLINK v1.9^63^. The resulting high-quality genotypes in PLINK format were then converted to VCF format for the imputation.

The genotype imputation for the RYSA and KOBS cohorts was separately performed on the Michigan imputation server against the HRC reference panel version r1.1 2016^64^ after removing SNPs with allele mismatch with the reference panel. On the server, haplotype phasing was performed using Eagle v2.4^65^ and the imputation was done using Minimac4^66^. The imputed genotype data were filtered by removing SNPs with imputation score r^2^<0.3 and HWE *p*-value<10^-6^.

For the unrelated METSIM participants, we performed quality control (QC) on the existing genotype data^26,59^ generated using an Illumina HumanOmniExpress BeadChip (Illumina), as described above. The genotype imputation was performed on the TOPMed imputation server against the TOPMed reference panel version r2^67^ using Minimac4^66^. Haplotype phasing was performed prior to the imputation using Eagle v2.4^65^ by the server. We performed QC on the imputed genotype data as described above.

#### Sample preparation and nuclei isolation for snRNA-seq of human SAT biopsies

To prepare for snRNA-seq experiments on the frozen SAT biopsies from the RYSA cohort, we designed a total of 21 batches with a pool of 8 samples per batch using random assignments. To minimize correlations between batches and phenotypic traits that include age, sex, BMI, and type 2 diabetes diagnosis status, we performed the random assignment of samples to batches multiple times and selected the set that resulted in a minimal correlation by batch. As we planned to demultiplex the pooled samples using individuals’ variant information, longitudinal samples from the same individual were each assigned to different batches. Based on the batch design, we pooled approximately 100 mg of each biopsy and isolated nuclei for 10x snRNA-seq, as previously described^42^. The nuclei concentration and overall quality were determined using Countess II after staining a small aliquot of nuclei from each batch with trypan blue and Hoechst dyes. We then immediate loaded the nuclei in a single channel of the 10x Chromium Chip for each batch and used the Single Cell 3’ v3.1 chemistry for library construction. The quality of cDNA and gene expression libraries were analyzed using an Agilent Bioanalyzer and the libraries from all batches were sequenced together on the NovaSeq S4 (Illumina) with a target sequencing depth of 400 million read pairs per batch.

#### Processing of human adipose snRNA-seq data

We processed the batches multiplexed of the human adipose snRNA-seq samples from RYSA by aligning the raw FASTQ format files against the GRCh38 human genome reference and GENCODE v42 annotations^68^ using STAR v2.7.10b^69^. Since we performed snRNA-seq, we used the ‘--soloFeatures GeneFull’ option in STAR to account for full pre-mRNA transcripts. The raw FASTQ files and the mapped BAM files from STAR alignment were then examined for their quality using FastQC.

#### Quality control and demultiplexing of the SAT snRNA-seq data

We developed a comprehensive QC pipeline, based on the QC of previous SAT snRNA-seq data cohorts by us and others^8,42^. First, we used DIEM v2.4.0^70^ to remove empty droplets and droplets with high amounts of ambient RNA. In DIEM, we used a UMI cutoff of 500 to set droplets below this cutoff as debris and increased the number of clusters for the initialization step with k-means clustering to 50 to separate droplets with high amounts of ambient RNA from true cell-type clusters. The default parameters were used for the remaining steps.

After running DIEM, we used Seurat v4.3.0.1^71^ to further remove low-quality droplets with UMIs≥500, nFeatures≥200, %mito≥10, and spliced RNA≥75%, given that a high unspliced RNA content is expected in nuclei. Then, we log-normalized the gene counts using the default scaling factor of 10,000. Next, top 2000 variable genes were calculated using the ‘FindVariableFeatures’ excluding mitochondrial and ribosomal genes, the normalized gene counts were scaled to mean 0 and variance 1, and the first 30 principal components (PCs) were calculated for clustering with a standard Louvain algorithm and a resolution of 0.5.

To further remove the possible remaining ambient RNA molecules, we used DecontX^72^ from celda R package v1.14.2 with Seurat cluster assignment as the ‘z’ and the previously removed low-quality droplets as the background to remove contaminated counts in each individual droplet. With the remaining clean counts, UMIs, nFeatures, and %mito were recalculated, and droplets were further filtered out by UMIs≥500, nFeatures≥200, and %mito≥10. Additionally, droplets with UMIs≥30,000 were also removed as very high UMIs likely indicate doublets.

As nuclei from the samples were pooled before sequencing, we demultiplexed each batch using demuxlet v2 from popscle software tool^73^ to identify the participant from whom the droplet originates. Demuxlet was ran with default parameters, except for ‘--min-MQ 30’, and imputed genotyped data after QC. We excluded droplets assigned as doublets or ambiguous and used the best matching individual to identify the originating individual for each droplet assigned as a singlet.

Finally, we used DoubletFinder v2.0.3^74^ to identify and remove any remaining doublets (e.g., doublets formed by nuclei from same individual). For each batch, we performed pN-pK parameter sweeps on a subset of 10,000 droplets and selected pN of 0.25 and the most optimal pK value maximizing mean-variance normalized bimodality coefficient. Then, we calculated pANN with the selected pN and pK values to make a density plot for each batch by pANN values and to estimate the predicted number of doublets. These values selected in the subset of 10,000 droplets were then applied to the full data to identify and remove doublets in each batch. Taken together this QC pipeline resulted in 137,061 nuclei from 167 samples passing the QC.

#### Data integration and cell-type and subcell-type annotation of SAT snRNA-seq data

All remaining high-quality droplets in the batches were first merged using Seurat v4.3.0.1^71^ and lowly expressed genes were excluded, keeping only the genes with at least 3 raw counts in at least 3 droplets (cells hereinafter). We repeated the gene count normalization, variable gene selection, scaling, and principal component analysis (PCA) steps, as described above in the merged data. To account for technical differences in gene expression driven by batch, we used Harmony v1.0.3^75^ to integrate on batch and performed clustering using the reductions from Harmony with a resolution of 0.5.

To annotate cells with their cell-type, we first assigned the clusters as adipocytes, ASPCs, lymphoid cells, myeloid cells, or vascular cells using SingleR v1.8.1^76^ with the SAT single cell and snRNA-seq data from the previously published adipose tissue atlas by Emont et al. as a reference^8^. Additionally, cell-type marker genes of each cluster were identified using a Wilcoxon rank sum test from the ‘FindAllMarkers’ function in Seurat (Bonferroni adjusted *p*-value<0.05) with logFC.threshold=0.25 and min.pct=0.25, as described previously^77,78^, and compared against the marker genes from the adipose atlas^8^. We then subset the five broad cell-types into individual objects and performed variable gene selection (2000), scaling, PCA, Harmony integration on batch, and clustering (Harmony dimensions 20-30 and resolutions 0.2-0.3) again to identify subclusters. Marker genes for each subcluster were identified similarly as described above, and the subcell-type annotations were assigned for the subclusters using SingleR v1.8.1^76^ and by comparing subcluster marker genes against the adipose atlas^8^. We determined the final resolution and subclusters for every broad cell-type when we observed that 1) unique marker genes are present in all subcell-types, and 2) no subcluster is specific to a single donor. Finally, we carried the subcell-type annotations back to the full object.

#### Functional enrichment of SAT subcell-type marker genes

We tested for enrichments of the gene ontology^79^ biological processes for each set of the SAT subcell-type markers using WebGestaltR v0.4.6^80^. All genes expressed, as defined as minimum 3 raw counts in at least 3 cells, in the corresponding broad cell-type were included as the reference gene set. We considered an enrichment to be significant with the Benjamini-Hochberg corrected *p*-value<0.05.

#### Differential composition analysis between time-points

To assess changes in cell-type composition between time-points, we performed the Friedman test for every cell-type using the cell-type proportions calculated at each time-point. As the accuracy of cell-type proportions might be reduced by the small number of nuclei sequenced in a given cell-type, we calculated cell-type proportions only for those with ≥500 nuclei from each time-point, including adipocytes, ASPCs, endothelial cells, macrophages, NK cells, pericytes, and T cells. For the cell-types that showed significant (*p*-value<0.05) differences in cell-type proportions across time-points, we performed pairwise Wilcoxon tests between all possible time-point pairs. We defined a significant difference in cell-type proportions between a pair of two time-points using the Bonferroni adjusted *p*-value<0.05.

#### Cell-type level pseudobulk differential expression analysis between time-points

We performed a cell-type level pseudobulk differential expression (DE) analysis for cell-types present in ≥5 individuals with ≥5 cells in each time-point. To do the analysis between the time-points, the 33 individuals with SAT snRNA-seq data at all four time-points were included in the time-point DE analysis. We first aggregated the raw counts of the genes with ≥3 raw counts in ≥3 cells^72^ for each snRNA-seq sample to obtain cell-type level pseudocounts. Next, we TMM normalized the expression values and used the limma v3.50.3^81^ and the voom normalization^82^, while correcting for age, sex, and number of nuclei as covariates. Only the expressed genes defined as having ≥1 pseudocount in ≥10% of samples in the smallest time-point group were included. As we have repeated measures data from the same 33 individuals across the four time-points, we used the ‘duplicateCorrelation’ function to account for the within individual correlation structure. The DE testing was done for 4 different contrasts to compare gene expression between the time-points a) 0m *vs* op, b) op *vs* 6m, c) 6m *vs* 12m, and d) op *vs* 12m. We defined a gene to be DE between time-points at the cell-type level using the Benjamini-Hochberg corrected *p*-value<0.05 and magnitude of logFC>0. Next, we tested for enrichments of the gene ontology^79^ biological processes separately for each up- and down-regulated cell-type level DEGs using Webgestalt^80^, as described above. We used all expressed genes tested for DE in each corresponding cell-type as the reference gene set and considered an enrichment to be significant with the FDR corrected *p*-value<0.05.

#### Comparison of SAT subcell-types with the white adipose tissue single cell atlas

As SAT subcell-types have been identified in the white adipose tissue single cell atlas by Emont et al.^8^, we sought to compare the subcell-types identified in the RYSA bariatric surgery cohort with those from the atlas. We downloaded the human SAT single cell RNA-seq data from Emont et al. and projected the subcell-type annotations to the RYSA SAT snRNA-seq data within each broad cell-type using Seurat v4.3.0.1^71^. First, we found anchor genes between the two cohorts using the ‘FindTransferAnchors’ function with the first 30 dimensions and the atlas data set as the reference and our data set as the query. Then, we used the ‘TransferData’ function with the anchor genes from the previous steps to assign subcell-type annotations from the atlas data to each of the subcell-type in the RYSA SAT snRNA-seq data.

#### Estimation of global association in adipocytes by change in BMI

To test for the heterogeneity in adipocytes based on individuals’ weight loss outcome, measured by the change in BMI (dBMI) between the op and 12m time-points and then adjusted by the BMI at the op time-point, we used the R package version of the co-varying neighborhood analysis (CNA) tool v0.0.99^11^. We employed the ‘association.Seurat’ function with 10,000 permutations and otherwise default parameters on the nuclei from the 33 individuals sampled at all four time-points that have been re-clustered and harmonized^75^ after subsetting for adipocytes. The contrast variable for the association test was set to dBMI and the covariates included were age, sex, and number of nuclei. We considered that the global association in adipocytes by dBMI is significant using global *p*-value<0.05 and that the neighborhood-level association in adipocytes by dBMI is significant using FDR<0.05.

#### Comparison of adipocyte and subcell-type proportions with BMI

We assessed the relationship between the adipocyte subcell-type proportions at each time-point with BMI. We first adjusted the adipocyte subcell-type proportions at each time-point for age and sex of individuals. Only the adipocyte subcell-types present in more than 20 individuals were included for each time-point. Next, we performed Spearman’s rank correlations on the age and sex adjusted proportions with BMI at the corresponding time-points. We defined a significant correlation using the Bonferroni adjusted *p*-value<0.05.

#### Cell-type level weighted gene co-expression network analysis

To identify cell-type level gene co-expression networks underlying adipocytes from each time-point, we used high dimensional weighted gene co-expression network analysis (hdWGCNA) v0.2.26^12^. We first employed the ‘MetacellsByGroups’ function with *k*=25 and the batch corrected Harmony reductions to aggregate transcriptionally similar cells within adipocytes at the operation time-point for constructing metacells, which improve the accuracy of the downstream co-expression network analysis by reducing sparsity and noise in the snRNA-seq data while retaining cellular heterogeneity. We limited our analysis to the cell-type expressed genes defined as ≥3 counts in ≥3 cells^72^ and created gene expression matrix using the constructed metacells. Next, we used the ‘TestSoftPowers’ function with ‘networkType=signed’ to compute the scale-free topology model fit for various softer power and then constructed a signed co-expression network in each broad cell-type using the ‘ConstructNetwork’ function with default parameters and a soft power automatically selected based on the parameter sweep from the previous step. We constructed the co-expression network in adipocytes from the op time-point as the accuracy of gene-gene correlations depends on the sparsity of the data and the number of cells was largest at op. The cell-type level co-expression networks from op were then projected onto the metacells from 12m time-point using the ‘ProjectModules’ function. For all co-expression networks, we computed module eigengenes (MEs) by performing PCA and taking the first PC within every network followed by Harmony batch correction. Each ME was harmonized by the sample of origin using the ‘ModuleEigengenes’ function. Lastly, we determined the eigengene-based connectivity (kME) of each gene using the ‘ModuleConnectivity’ on the full snRNA-seq data to identify top hub genes in the networks. To find functional enrichments, we performed the gene ontology^79^ biological processes enrichment analysis on the network genes using Webgestalt^80^, as described above.

We sought to identify networks enriched for adipocyte time-point DEGs and subcell-type unique marker genes by running the Fisher’s exact test on network genes overlapping the time-point DE or subcell-type unique marker gene sets as implemented by the ‘OverlapModulesDEGs’ function. We considered a network to be significantly enriched for a gene set with an FDR corrected Fisher’s *p*-value<0.05.

We performed network preservation analysis of the op time-point networks in the 33 individuals’ 12m time-point SAT snRNA-seq data, and in independent SAT snRNA-seq data from the 35 individuals’ op time-point biopsies as replication. These analyses were conducted using the ‘ModulePreservation’ function in hdWGNCA^12^, and a network preservation Z summary score was employed to consider a network to be highly preserved (Z>10), moderately preserved (2≤Z≤10), or not preserved (Z<2)^83^.

#### GWAS gene set enrichment analysis with MAGMA

To search for enrichment of BMI GWAS signals in the cell-type co-expression networks, we used MAGMA (Multi-marker Analysis of GenoMic Annotation) v1.10^13^. We used the publicly available BMI GWAS summary statistics from FinnGen^84^ consortium (version R10) with the METSIM cohort genotype data as the LD reference.

#### Processing of human preadipocyte and adipocyte bulk assay for transposase accessible chromatin (ATAC)-seq data

We used bulk ATAC-seq data previously generated from human primary adipocytes after 14-day differentiation of preadipocytes to identify open chromatin regions in adipocytes^21^. The preadipocyte differentiation experiment was conducted as described in detail previously^21,34^. We followed the Omni-ATAC protocol^85^ to generate the ATAC-seq data with an average of 126M (SD=48M) reads per sample. We performed QC on the data following the Omni-ATAC protocol^85^, as described previously^86^ and used MACS2 v2.2.7.1 to call peaks while removing the peaks in the blacklisted regions. The merged peaks across the time-points generated the final consensus peak set of 135,190 peaks.

#### ATAC-peak stratified partitioned heritability analysis

We conducted a partitioned heritability analysis using stratified LD-score regression (LDSC) v1.0.1^14^ to investigate whether the variants residing in adipocyte open chromatin regions are enriched for the heritability of BMI and WHRadjBMI. Briefly, we estimated LD scores with LDSC v1.0.1^14^ from the following five annotations: all SNPs in the genome, as well as SNP residing in the adipocyte peaks and outside of the adipocyte peaks, respectively. All annotations were limited to SNPs with MAF>5%^87^. We used the publicly available BMI GWAS summary statistics from FinnGen^84^ consortium (version R10) and the WHRadjBMI GWAS summary statistics from the GIANT-UKB meta-analysis^88^ to provide the variant-level weights. Then we computed the BMI and WHRadjBMI heritability per each annotation with LDSC v1.0.1^14^ and tested each peak annotation for heritability enrichment, i.e., the proportion of heritability divided by the proportion of the genome from the annotation^14^.

#### Partitioned heritability analysis of adipocyte network genes

To assess whether the MAGMA enriched adipocyte network gene sets harbored SNPs in their *cis* regions (±500kb of gene body) enriched for BMI and WHRadjBMI heritability, we performed partitioned heritability analysis using LDSC v1.0.1^14^. For each network gene set, we estimated LD scores from all SNPs with MAF>0.05 within the *cis* regions (±500kb from the ends) of the network genes and of the weight loss DEGs in the network, computed the BMI and WHRadjBMI heritability per annotation using the GWAS summary statistics, and tested the heritabilities for enrichments. We then repeated the analysis by subsetting SNPs in *cis* regions to those also residing in the adipocyte open chromatin peaks. For all LDSC analyses, only the SNP sets covering at least 1% of the genome were tested due to the limited power of heritability estimators for small annotations, as recommended^14^.

#### Cell-type level *cis*-eQTL analysis of human SAT snRNA-seq data at each time-point

We performed cell-type level pseudobulk *cis*-eQTL analysis in the 33 individuals with SAT snRNA-seq data from all four time-points by aggregating the raw counts of genes measured in adipocytes for the samples at the four time-points to construct pseudocount matrices. We limited the testing to imputed and QC passing SNPs with MAF>10% in the larger Finnish KOBS (n=509) cohort, which resulted in 4,248,579 SNPs, and to genes with pseudocount thresholds of ≥0.1 transcripts per million (TPM) in ≥20% of samples and ≥6 raw reads in ≥20% of samples^22^. The pseudocount expression values for each gene were then normalized between samples using TMM^89^ and inverse normal transformed across samples. Next, we performed PCA on the gene expression matrices and selected optimal number of PCs to include as covariates using the Buja and Eyuboglu (BE) algorithm^90^, implemented in PCAforQTL v0.1.0^91^ to capture hidden confounders of expression variability. We performed *cis*-eQTL analysis using Matrix eQTL v2.3^92^ while including top expression PCs and number of nuclei as covariates. To compare the results across time-points, we limited these analyses to the same SNP-gene pairs in all four time-points, which included 13,415 genes for adipocytes. We defined *cis* regions as ±500kb from the ends of genes and significant *cis*-eQTLs using an FDR<0.05.

To search for shared- and time-point-specific *cis*-eQTLs, we jointly analyzed the *cis*-eQTL effects across all time-points using the multivariate adaptive shrinkage software in R (mashr v0.2.50)^93^. We first performed time-point-by-time-point analysis using the ‘mash_1by1’ function after pruning highly correlated SNPs (--indep-pairwise 250 50 0.9)^24^ using PLINK v1.9^63^. For each gene, we then used the top associated SNPs that had local false sign rate (LFSR)<0.1 and used these top associated SNPs to estimate data-driven covariance matrices. The default canonical covariance matrices were estimated in a random subset of 20,000 SNP-gene pairs. Next, we fitted the mash model to the random 20,000 SNP-gene pair subset with the data-driven and canonical covariances, and then computed the posterior summaries on all SNP-gene pairs across all time-points using the mash fit. We defined an eGene to be time-point-shared if it had at least one significant *cis*-eQTL SNP with LFSR<0.05 in all four time-points, and to be time-point-specific if it had a *cis*-eQTL SNP (LFSR<0.05) only in one time-point.

#### Replication of adipocyte *cis*-eQTLs among the bulk SAT *cis*-eQTLs

We compared the adipocyte pseudobulk *cis*-eQTLs with bulk SAT *cis*-eQTLs from previously published Finnish bariatric surgery cohort, KOBS (n=260)^21^, and GTEx^22^. To minimize the *cis-* eQTL method based differences in the replication analysis, we used Matrix eQTL v2.3^92^ to perform *cis*-eQTL analysis in the GTEx v8 bulk SAT RNA-seq data (n=581). We included top 5 genotype PCs, sequencing platform, sequencing protocol, sex, and top 60 Probabilistic Estimation of Expression Residuals (PEER) factors as covariates and tested for variants with MAF>0.01 and highly expressed genes as in the original publication^22^. For the replication analysis in the bulk SAT RNA-seq cohorts, we further restricted the SNP-gene pairs to those that were significant (LFSR<0.05) in our adipocyte pseudobulk *cis*-eQTL analysis at the operation time-point and performed a *q*-value correction using a *q*-value threshold of 0.05. We first checked for the replication of eGenes with at least one shared *cis*-eQTL SNP in the same direction of effect between the adipocyte and bulk SAT *cis*-eQTL cohorts. Next, we checked for the replication of the adipocyte lead SNP-gene pairs in the bulk SAT *cis*-eQTL cohorts. Finally, we calculated the ν_1_ statistic^23^ to estimate the proportion of the adipocyte lead SNP-gene pairs that replicated in the bulk SAT *cis*-eQTL cohorts.

#### Subcell-type level *cis*-eQTL analysis of human adipocytes at each time-point

We performed subcell-type level pseudobulk *cis*-eQTL analysis for the adipocyte subcell-types, Ad1 and Ad3, from all four time-points by aggregating the raw counts of genes measured in each subcell-type to construct pseudocount matrices, as described above. As in the cell-type level pseudobulk *cis*-eQTL analysis, we constructed pseudocount matrices only for the subcell-types from individuals with ≥5 nuclei^24^; however, we allowed for less than 33 individuals to have ≥5 nuclei (n=31, 32, 32, and 31 individuals for Ad1 and n=21, 31, 29, and 27 individuals for Ad3 at 0m, op, 6m, and 12m time-points, respectively), considering that subcell-types have lower numbers of nuclei than the main cell-types. The same SNP and gene filtering criteria were used, which resulted in 10,818 and 2,661 genes tested for Ad1 and Ad3, respectively. We performed *cis*-eQTL analysis using Matrix eQTL v2.3^92^, followed by shared- and time-point-specific *cis*-eQTL analysis using mashr v0.2.50^93^, as described above.

#### Construction of genome-wide polygenic risk scores for BMI

To construct polygenic risk scores (PRSs) for BMI for the individuals in the METSIM (n=8,254), KOBS (n=495), and RYSA (n=68) cohorts, we used the publicly available BMI GWAS summary statistics from the FinnGen consortium^84^ (version R10) as the base data for the SNP effect weights, and the METSIM genotype data^26^ as the target data for building the PRS model. We followed the QC pipeline from^94^ with minor modifications. Briefly, we removed SNPs with MAF<1% and strand ambiguous SNPs from the base data, and SNPs with MAF<5%, imputation r^2^<0.3, SNP call rate<99%, Hardy Weinberg *p*-value<10^−6^, or not present in the base data from the genotype data^94^ using PLINK v1.9^63^. Individuals with up to second degree relatedness, as determined by KING v2.3.2^95^, sex discordance, genotype call rate<98%, or heterozygosity above 4 SD from the mean were also removed^84,94^ using PLINK v1.9^63^. To build the genome-wide PRS model from all variants landing in the genome, we first obtained independent variants by performing LD-clumping on the target data SNPs using PLINK v1.9^63^, with r^2^<0.1, and a window size of 250kb. We empirically determined the optimal p-value threshold by constructing PRSs in the target data from the clumped SNPs with a range of *p*-value thresholds (5×10^−8^, 1×10^−7^, 1×10^−6^, 1×10^−5^, 1×10^−4^, 5×10^−4^, 0.001, 0.005, 0.01, 0.05, 0.1, 0.2, 0.3, 0.4, 0.5), and identifying the PRS with the highest R^2^ for BMI relative to a null model containing age and 10 genetic PCs as the covariates, and quantile normalized BMI as the outcome^94^. The genome-wide PRSs were then computed together from the genotype data of METSIM, KOBS and RYSA from the METSIM clumped SNPs with the optimal *p*-value threshold (*p*-value<0.4).

#### Prediction of weight loss using polygenic risk scores for BMI

We first constructed two partitioned network BMI PRS sets in the independent Finnish bariatric surgery cohort, KOBS using the *cis*-eQTL SNPs of the DEGs in the Ad network VIII and Ad network VIII further subset for DEGs upregulated at the op time-point in RYSA that also land in adipocyte open chromatin regions. We employed the set-based clumping method of PRSet implemented in PRSice v2.3.5^96,97^ for these analyses. As in the genome-wide BMI PRS, we used the METSIM genotype data to LD-clump variants with an r^2^ cutoff of 0.1 and window of 250kb, and BMI GWAS summary statistics (version R10) from FinnGen^84^ to provide the model weights.

Next, we used the genome-wide BMI PRS and these two network BMI PRS sets constructed in the KOBS cohort to predict the weight loss outcome, defined by BMI change between the operation and 12-month time-points and adjusted by BMI at the operation time-point (dBMI). We restricted our analysis to the 170 individuals of the KOBS cohort with both the operation and 12-month follow-up BMI measurements available to be able to calculate the dBMI values. We fitted a linear model with age, sex, 10 genetic PCs, and baseline BMI as covariates and quantile normalized dBMI as the outcome. We performed an F-test to assess the model for improved fit over a reduced model consisting of only covariates. For the network PRS sets with significant model improvement (*p*-value<0.05), we then derived competitive *p*-values with PRSet by ranking the R^2^ of BMI change for the network PRS relative to that of 10,000 random PRSs (i.e., 10,000 permutations), each built from a randomly selected independent set of SNPs of the same size from the *cis* regions (±500kb from the ends of gene bodies) of cell-type expressed genes. We considered a PRS for BMI to be a significant predictor of weight loss with *p*_perm10,000_<0.05.

## Code availability

No custom code was used, and all codes used for analyses in this study were based on the publicly available source code of the packages listed in the Methods.

## Data Availability

The SAT snRNA-seq data will be made available in the NIH Gene Expression Omnibus (GEO) upon acceptance, under accession number GSEXX. The cell-type level *cis*-eQTL summary level data will be made available in the UCLA Dataverse, at https://dataverse.ucla.edu/XX. The adipocyte ATAC-seq data are available in GEO, under accession number GSE269929^21^. Data from the UK Biobank were used in this study under UK Biobank Application Number 33934. UK Biobank data are available for bona fide researchers through the application process (https://www.ukbiobank.ac.uk/learn-more-about-uk-biobank/contact-us). The GTEx whole-genome sequencing and gene expression data are available at dbGaP phs000424.v8.p2.

## Acknowledgements

We thank the participants of the RYSA, KOBS, and UK Biobank studies. This research was conducted using the UK Biobank Resource under application number 33934. This work uses data provided by patients and collected by the NHS as part of their care and support. The GTEx datasets used for the analyses described in the present study were obtained from dbGaP at http://www.ncbi.nlm.nih.gov/gap through dbGaP accession phs000424.v8.p2.

## Author contributions

S.H.T.L. and P.P. designed the study. S.H.T.L., A.K., M.A., B.B., and P.P. developed the methods and statistical approaches. S.H.T.L., A.K., and K.Z.G performed computational and statistical analyses. S.H.T.L and S.R. generated the RYSA SAT snRNA-seq data. M.A. generated the RYSA genotype data. S.H., B.v.d.K., K.H.P, U.S., T.S., and A.J. collected the RYSA clinical data and samples. D.K., P.P., and J.P. generated the KOBS genotype data. V.M. and J.P. collected the KOBS clinical data and samples. M.L. generated the METSIM genotype data. K.M.G. and P.P. performed the human primary preadipocyte differentiation experiment and generated the ATAC-seq data. S.H.T.L and P.P wrote the manuscript, and all authors read, reviewed, or edited the manuscript. All authors read and approved the final manuscript.

---

**Supplementary Figure 1.**
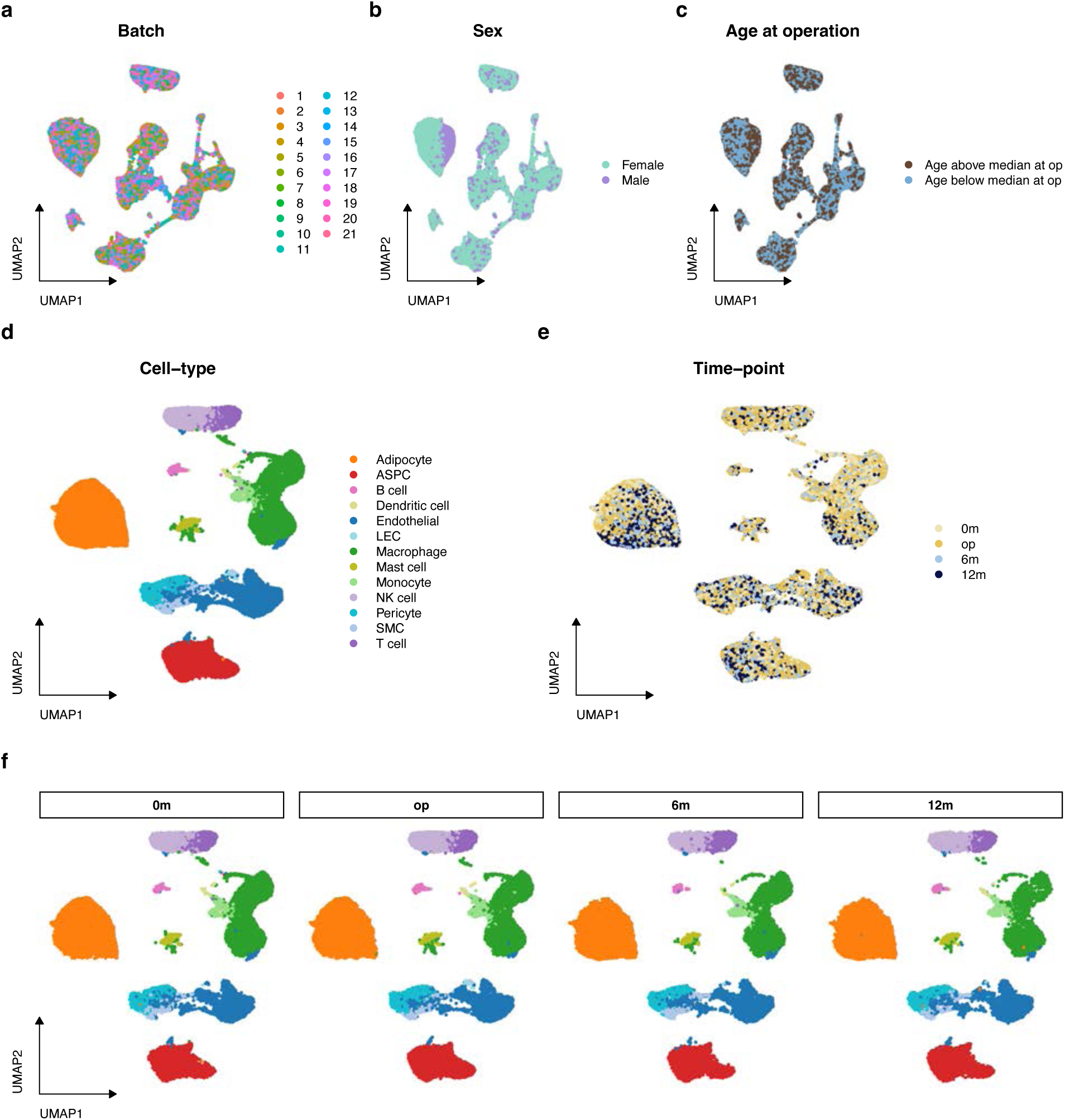
Construction of longitudinal SAT snRNA-seq data. **a–c** Uniform Manifold Approximation and Projection (UMAP) visualization of 137,061 nuclei collected from 167 SAT biopsies across four time-points (0m, baseline; op, operation; 6m, 6 months after operation; and 12m, 12 months after operation), colored by the originating experiment batch **(a)**, sex **(b)** of individuals, and age at the operation time-point **(c)**. **d–f** UMAP visualization of a subset of the nuclei (n=104,995 nuclei from 132 SAT biopsies), originating from the 33 individuals who had SAT biopsies collected from all four time-points, colored by the assigned cell-type **(d)**, originating biopsy time-point **(e)**, and separated by the originating biopsy time-point **(f)**. ASPC indicates adipose stem and progenitor cells; LEC, lymphatic endothelial cells; NK, natural killer cells; and SMC, smooth muscle cells.

**Supplementary Figure 2.**
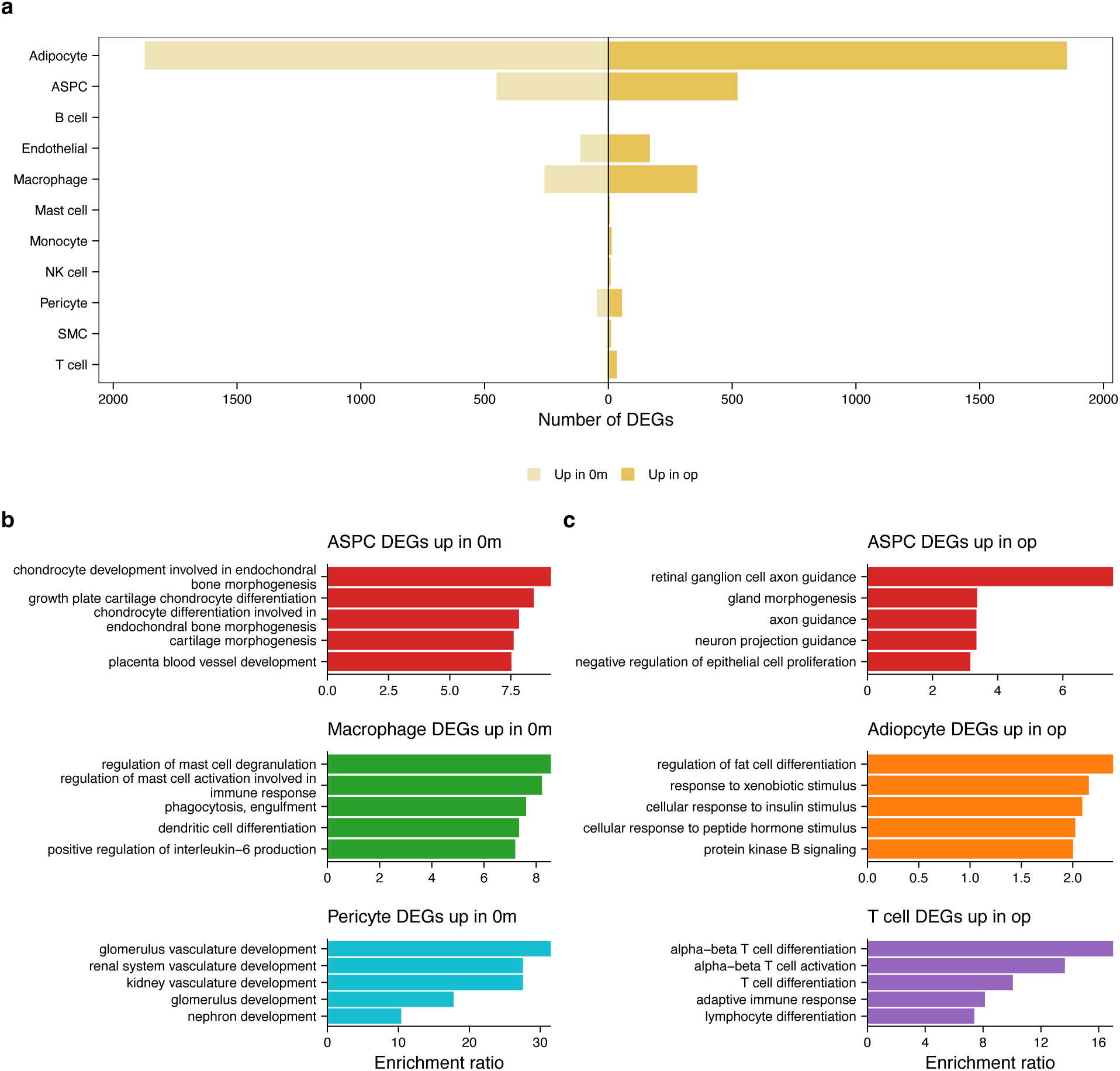
Adipocytes exhibit the largest change in gene expression after the very low-caloric diet. **a** Number of differentially expressed genes (FDR<0.05) between the 0-month (0m) and operation (op) time-points per cell-type are shown colored by the time-point, in which the genes are upregulated. **b** Top five gene ontology (GO) biological process terms by the genes upregulated at the 0m time-point in ASPCs, macrophages, and pericytes **(b)** and by the genes upregulated at the op time-point in ASPCs, adipocytes, and T cells **(c)**. ASPC indicates adipose stem and progenitor cells; DEGs, differentially expressed genes; NK cell, natural killer cells, and SMC, smooth muscle cells.

**Supplementary Figure 3.**
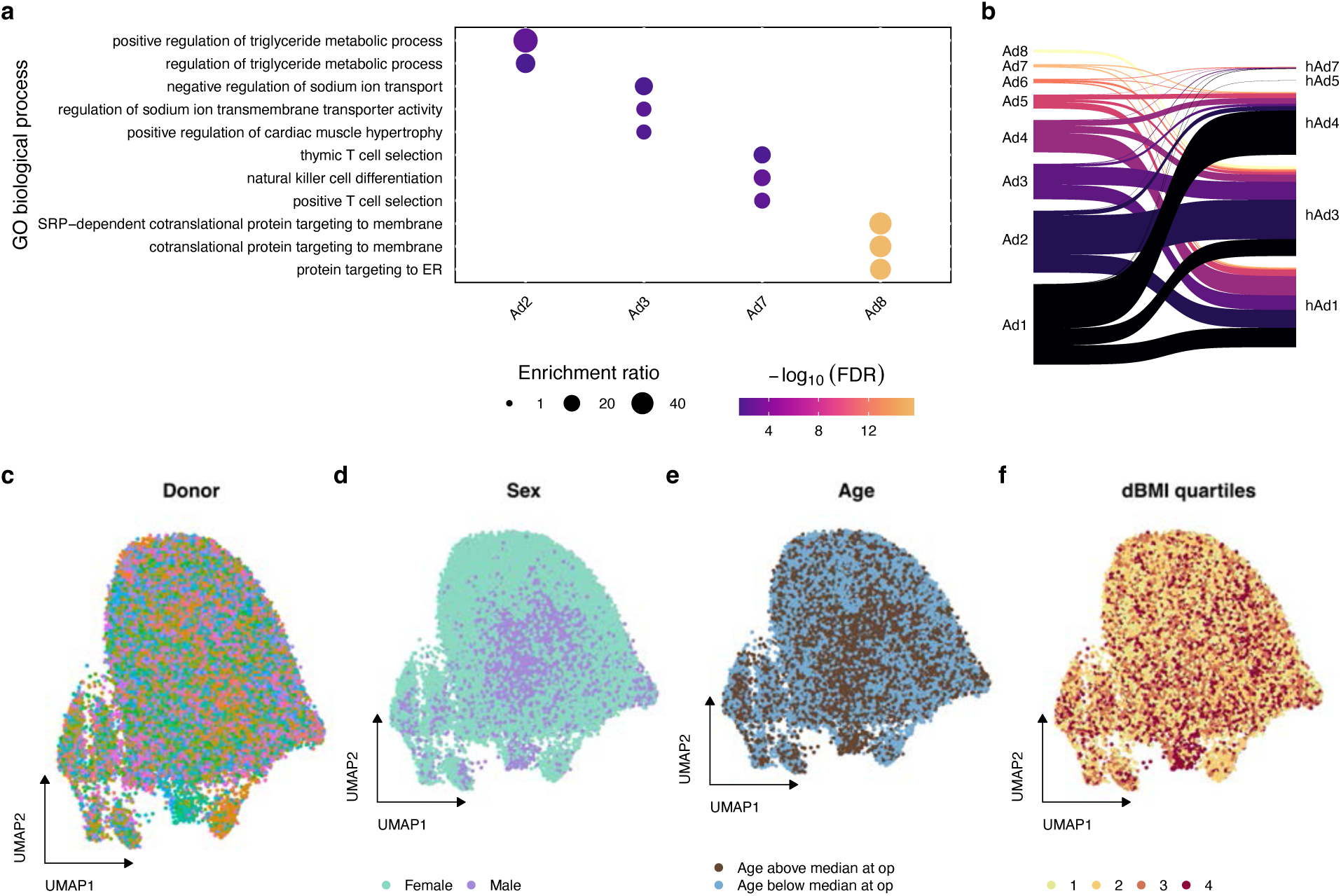
Adipocyte subcell-type composition reveals heterogeneity. **a** Dot plot showing enrichment of the top three gene ontology (GO) biological process terms by the unique adipocyte subcell-type marker genes based on the significant (FDR<0.05) enrichment ratio. The dots are colored by significance of the enrichment (–log_10_FDR) and their size represents the enrichment ratio. **b** River plot showing the correspondence between the adipocyte subcell-types classified in this study and the subcell-type annotations from the previous adipose single-cell atlas^1^. The subcell-type annotations from this study are shown on the left and the those from the previous adipose atlas^1^ on the right. **c–f** Uniform Manifold Approximation and Projection (UMAP) visualization of adipocytes collected from the 33 individuals across four time-points (0m, baseline; op, operation; 6m, 6 months after operation; and 12m, 12 months after operation) (n=132 SAT biopsies), colored by the originating individuals **(c)**, sex **(d)** of individuals, age at the operation time-point **(e)**, and change in BMI between the op and 12m time-points (dBMI) **(f)** grouped into quartiles, where the largest value represents the top 25% of the individuals who observed the greatest weight loss after bariatric surgery. Ad indicates adipocytes and FDR, false discovery rate.

**Supplementary Figure 4.**
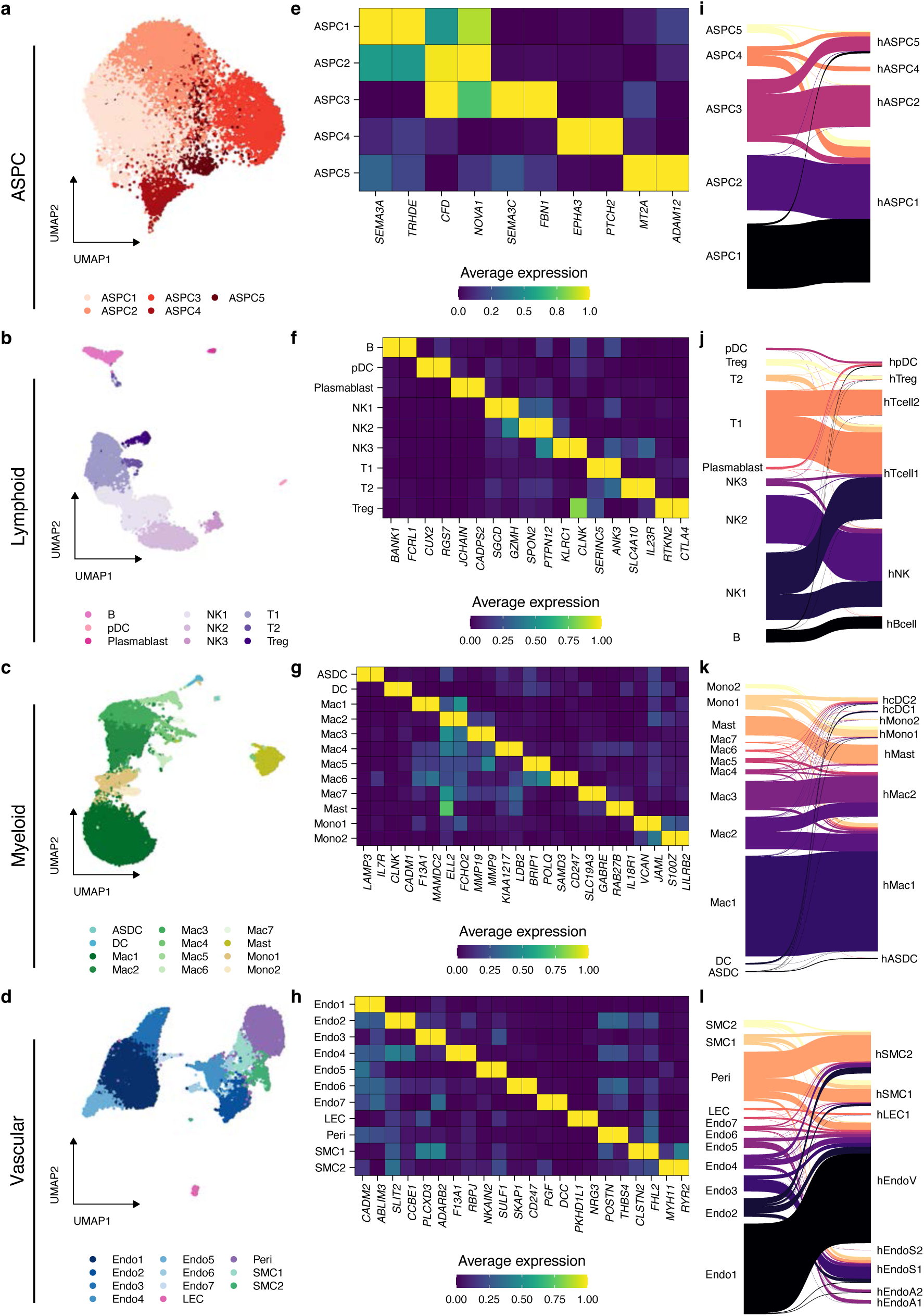
Identification of subcell-types in SAT. **a–d** Uniform Manifold Approximation and Projection (UMAP) visualization of ASPC (**a**), lymphoid **(b)**, myeloid **(c)**, and vascular **(d)** cells collected from the 33 individuals with SAT snRNA-seq data available at four time-points (0m, baseline; op, operation; 6m, 6 months after operation; and 12m, 12 months after operation) (n=132 SAT biopsies), colored by subclusters. **e–h** Average normalized expression of the top protein coding subcell-type marker genes used to annotate each subcell-type in ASPC **(e)**, lymphoid **(f)**, myeloid **(g)**, and vascular **(h)** cells. **i–l** River plot showing the correspondence between the subcell-types classified in this study and the subcell-type annotations from the previous adipose single-cell atlas^8^ for ASPC **(i)**, lymphoid **(j)**, myeloid **(k)**, and vascular **(l)** cells. The subcell-type annotations from this study are shown on the left and the those from the previous adipose atlas^1^ on the right. ASPC indicates adipose stem and progenitor cells and SAT, subcutaneous adipose tissue.

**Supplementary Figure 5.**
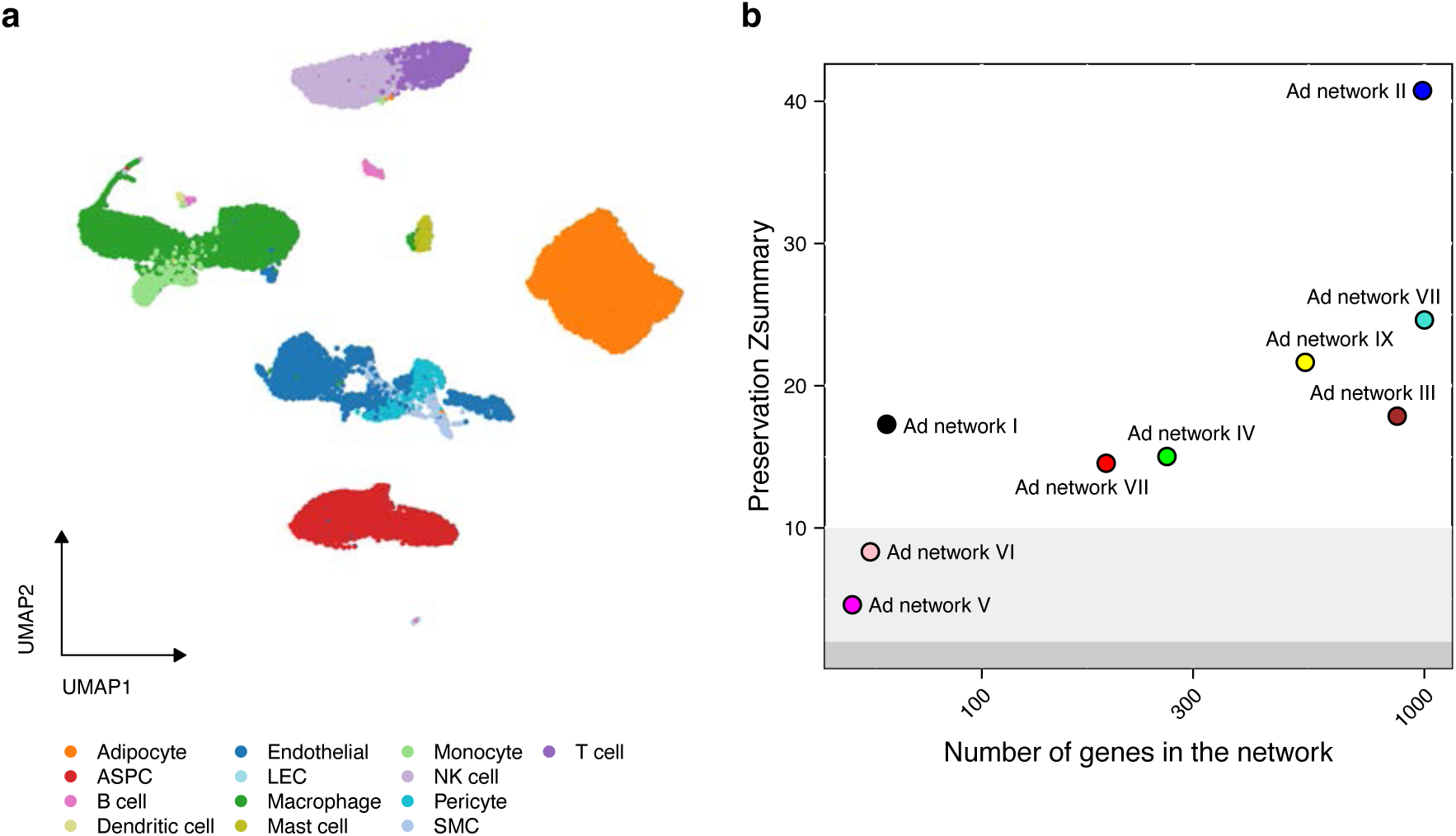
Cell-type level networks are largely replicated. **a** Uniform Manifold Approximation and Projection (UMAP) visualization of 32,066 nuclei collected from the 35 individuals with SAT biopsies at the operation (op) time-point colored by the assigned cell-type. **b** The preservation Z summary scores computed using hdWGCNA^2^ for the cell-type level networks (n=33 individuals), included in the main analyses at the operation (op) time-point, are shown in the replication dataset of 35 individuals at the op time-point, plotted by the size of each network for adipocytes. The background of the scatterplot is shaded based on the preservation of the networks, where dark gray (Z≤2) indicates not preserved, light gray (2<Z≤10) moderately preserved, and no shade (Z>10) highly preserved, respectively. ASPC indicates adipose stem and progenitor cells; LEC, lymphatic endothelial cells; NK, natural killer cells; and SMC, smooth muscle cells.

## References

1. Valenzuela, P. L. et al. Obesity and the risk of cardiometabolic diseases. Nat. Rev. Cardiol. 20, 475–494 (2023).

2. Khera, A. V. et al. Polygenic Prediction of Weight and Obesity Trajectories from Birth to Adulthood. Cell 177, 587–596.e9 (2019).

3. O’Brien, P. E. et al. Long-Term Outcomes After Bariatric Surgery: a Systematic Review and Meta-analysis of Weight Loss at 10 or More Years for All Bariatric Procedures and a Single-Centre Review of 20-Year Outcomes After Adjustable Gastric Banding. Obes. Surg. 29, 3– 14 (2019).

4. Dent, R., McPherson, R. & Harper, M.-E. Factors affecting weight loss variability in obesity. Metabolism. 113, 154388 (2020).

5. Pellegrinelli, V., Carobbio, S. & Vidal-Puig, A. Adipose tissue plasticity: how fat depots respond differently to pathophysiological cues. Diabetologia 59, 1075–1088 (2016).

6. Sakers, A., De Siqueira, M. K., Seale, P. & Villanueva, C. J. Adipose-tissue plasticity in health and disease. Cell 185, 419–446 (2022).

7. Cruz-García, E. M., Frigolet, M. E., Canizales-Quinteros, S. & Gutiérrez-Aguilar, R. Differential Gene Expression of Subcutaneous Adipose Tissue among Lean, Obese, and after RYGB (Different Timepoints): Systematic Review and Analysis. Nutrients 14, 4925 (2022).

8. Emont, M. P. et al. A single-cell atlas of human and mouse white adipose tissue. Nature 603, 926–933 (2022).

9. Heinonen, S. et al. Roux-en-Y versus one-anastomosis gastric bypass (RYSA study): weight loss, metabolic improvements, and nutrition at 1 year after surgery, a multicenter randomized controlled trial. Obes. Silver Spring Md 31, 2909–2923 (2023).

10. Li, S.-N. & Wu, J.-F. TGF-β/SMAD signaling regulation of mesenchymal stem cells in adipocyte commitment. Stem Cell Res. Ther. 11, 41 (2020).

11. Reshef, Y. A. et al. Co-varying neighborhood analysis identifies cell populations associated with phenotypes of interest from single-cell transcriptomics. Nat. Biotechnol. 40, 355–363 (2022).

12. Morabito, S., Reese, F., Rahimzadeh, N., Miyoshi, E. & Swarup, V. hdWGCNA identifies co-expression networks in high-dimensional transcriptomics data. *Cell Rep*. Methods 3, 100498 (2023).

13. de Leeuw, C. A., Mooij, J. M., Heskes, T. & Posthuma, D. MAGMA: generalized gene-set analysis of GWAS data. PLoS Comput. Biol. 11, e1004219 (2015).

14. Finucane, H. K. et al. Partitioning heritability by functional annotation using genome-wide association summary statistics. Nat. Genet. 47, 1228–1235 (2015).

15. Emdin, C. A. et al. Genetic Association of Waist-to-Hip Ratio With Cardiometabolic Traits, Type 2 Diabetes, and Coronary Heart Disease. JAMA 317, 626 (2017).

16. Currin, K. W. et al. Genetic effects on liver chromatin accessibility identify disease regulatory variants. Am. J. Hum. Genet. 108, 1169–1189 (2021).

17. Perrin, H. J. et al. Chromatin accessibility and gene expression during adipocyte differentiation identify context-dependent effects at cardiometabolic GWAS loci. PLoS Genet. 17, e1009865 (2021).

18. Arthur, T. D. et al. Multiomic QTL mapping reveals phenotypic complexity of GWAS loci and prioritizes putative causal variants. Cell Genomics 100775 (2025) doi:10.1016/j.xgen.2025.100775.

19. Christodoulides, C., Lagathu, C., Sethi, J. K. & Vidal-Puig, A. Adipogenesis and WNT signalling. Trends Endocrinol. Metab. TEM 20, 16–24 (2009).

20. Jang, M. J. et al. CACUL1 reciprocally regulates SIRT1 and LSD1 to repress PPARγ and inhibit adipogenesis. Cell Death Dis. 8, 3201 (2017).

21. Lee, S. H. T. et al. Single nucleus RNA-sequencing integrated into risk variant colocalization discovers 17 cell-type-specific abdominal obesity genes for metabolic dysfunction-associated steatotic liver disease. EBioMedicine 106, 105232 (2024).

22. GTEx Consortium. The GTEx Consortium atlas of genetic regulatory effects across human tissues. Science 369, 1318–1330 (2020).

23. Fujita, M. et al. Cell subtype-specific effects of genetic variation in the Alzheimer’s disease brain. Nat. Genet. 56, 605–614 (2024).

24. Natri, H. M. et al. Cell-type-specific and disease-associated expression quantitative trait loci in the human lung. Nat. Genet. 56, 595–604 (2024).

25. Emani, P. S. et al. Single-cell genomics and regulatory networks for 388 human brains. Science 384, eadi5199 (2024).

26. Laakso, M. et al. The Metabolic Syndrome in Men study: a resource for studies of metabolic and cardiovascular diseases. J. Lipid Res. 58, 481–493 (2017).

27. Pihlajamäki, J. et al. Cholesterol absorption decreases after Roux-en-Y gastric bypass but not after gastric banding. Metabolism 59, 866–872 (2010).

28. Nica, A. C. et al. Candidate Causal Regulatory Effects by Integration of Expression QTLs with Complex Trait Genetic Associations. PLoS Genet. 6, e1000895 (2010).

29. Nicolae, D. L. et al. Trait-Associated SNPs Are More Likely to Be eQTLs: Annotation to Enhance Discovery from GWAS. PLoS Genet. 6, e1000888 (2010).

30. Torres, J. M. et al. Cross-tissue and tissue-specific eQTLs: partitioning the heritability of a complex trait. Am. J. Hum. Genet. 95, 521–534 (2014).

31. Dornbos, P. et al. Evaluating human genetic support for hypothesized metabolic disease genes. Cell Metab. 34, 661–666 (2022).

32. Kerr, A. G., Andersson, D. P., Rydén, M., Arner, P. & Dahlman, I. Long-term changes in adipose tissue gene expression following bariatric surgery. J. Intern. Med. 288, 219–233 (2020).

33. Mardinoglu, A. et al. Extensive weight loss reveals distinct gene expression changes in human subcutaneous and visceral adipose tissue. Sci. Rep. 5, 14841 (2015).

34. Kar, A. et al. Age-dependent genes in adipose stem and precursor cells affect regulation of fat cell differentiation and link aging to obesity via cellular and genetic interactions. Genome Med. 16, 19 (2024).

35. Repsilber, D. et al. Biomarker discovery in heterogeneous tissue samples-taking the in-silico deconfounding approach. BMC Bioinformatics 11, 27 (2010).

36. Glastonbury, C. A., Couto Alves, A., El-Sayed Moustafa, J. S. & Small, K. S. Cell-Type Heterogeneity in Adipose Tissue Is Associated with Complex Traits and Reveals Disease-Relevant Cell-Specific eQTLs. Am. J. Hum. Genet. 104, 1013–1024 (2019).

37. Vijay, J. et al. Single-cell analysis of human adipose tissue identifies depot- and disease-specific cell types. Nat. Metab. 2, 97–109 (2019).

38. Hildreth, A. D. et al. Single-cell sequencing of human white adipose tissue identifies new cell states in health and obesity. Nat. Immunol. 22, 639–653 (2021).

39. Massier, L. et al. An integrated single cell and spatial transcriptomic map of human white adipose tissue. Nat. Commun. 14, 1438 (2023).

40. Aron-Wisnewsky, J., Doré, J. & Clement, K. The importance of the gut microbiota after bariatric surgery. Nat. Rev. Gastroenterol. Hepatol. 9, 590–598 (2012).

41. Kong, L.-C. et al. Gut microbiota after gastric bypass in human obesity: increased richness and associations of bacterial genera with adipose tissue genes. Am. J. Clin. Nutr. 98, 16–24 (2013).

42. Pan, D. Z. et al. Identification of TBX15 as an adipose master trans regulator of abdominal obesity genes. Genome Med. 13, 123 (2021).

43. Raulerson, C. K. et al. Adipose Tissue Gene Expression Associations Reveal Hundreds of Candidate Genes for Cardiometabolic Traits. Am. J. Hum. Genet. 105, 773–787 (2019).

44. Fairfax, B. P. et al. Innate immune activity conditions the effect of regulatory variants upon monocyte gene expression. Science 343, 1246949 (2014).

45. Alasoo, K. et al. Shared genetic effects on chromatin and gene expression indicate a role for enhancer priming in immune response. Nat. Genet. 50, 424–431 (2018).

46. Soskic, B. et al. Immune disease risk variants regulate gene expression dynamics during CD4+ T cell activation. Nat. Genet. 54, 817–826 (2022).

47. Lass, A. et al. Adipose triglyceride lipase-mediated lipolysis of cellular fat stores is activated by CGI-58 and defective in Chanarin-Dorfman Syndrome. Cell Metab. 3, 309–319 (2006).

48. Redaelli, C. et al. Clinical and genetic characterization of Chanarin-Dorfman syndrome patients: first report of large deletions in the ABHD5 gene. Orphanet J. Rare Dis. 5, 33 (2010).

49. Locke, A. E. et al. Genetic studies of body mass index yield new insights for obesity biology. Nature 518, 197–206 (2015).

50. Delahanty, L. M. et al. Genetic predictors of weight loss and weight regain after intensive lifestyle modification, metformin treatment, or standard care in the Diabetes Prevention Program. Diabetes Care 35, 363–366 (2012).

51. Rosenbaum, M. & Foster, G. Differential mechanisms affecting weight loss and weight loss maintenance. Nat. Metab. 5, 1266–1274 (2023).

52. Peña, E. et al. Use of polygenic risk scores to assess weight loss after bariatric surgery: a five-year follow-up study. J. Gastrointest. Surg. Off. J. Soc. Surg. Aliment. Tract S1091–255X(24)00485–2 (2024) doi:10.1016/j.gassur.2024.05.029.

53. Pei, H., Yao, Y., Yang, Y., Liao, K. & Wu, J.-R. Krüppel-like factor KLF9 regulates PPARγ transactivation at the middle stage of adipogenesis. Cell Death Differ. 18, 315–327 (2011).

54. Buerger, F. et al. Depletion of Jmjd1c impairs adipogenesis in murine 3T3-L1 cells. Biochim. Biophys. Acta Mol. Basis Dis. 1863, 1709–1717 (2017).

55. Viscarra, J. A., Wang, Y., Nguyen, H. P., Choi, Y. G. & Sul, H. S. Histone demethylase JMJD1C is phosphorylated by mTOR to activate de novo lipogenesis. Nat. Commun. 11, 796 (2020).

56. Dairi, G. et al. Transcriptomic and Proteomic Analysis Reveals the Potential Role of RBMS1 in Adipogenesis and Adipocyte Metabolism. Int. J. Mol. Sci. 24, 11300 (2023).

57. Lima, M. M. O. et al. Visceral fat resection in humans: effect on insulin sensitivity, beta-cell function, adipokines, and inflammatory markers. Obes. Silver Spring Md 21, E182–189 (2013).

58. Cariou, B. The metabolic triad of non-alcoholic fatty liver disease, visceral adiposity and type 2 diabetes: Implications for treatment. Diabetes Obes. Metab. 24 **Suppl 2**, 15–27 (2022).

59. Pan, D. Z. et al. Integration of human adipocyte chromosomal interactions with adipose gene expression prioritizes obesity-related genes from GWAS. Nat. Commun. 9, 1512 (2018).

60. Littlejohns, T. J. et al. The UK Biobank imaging enhancement of 100,000 participants: rationale, data collection, management and future directions. Nat. Commun. 11, 2624 (2020).

61. Sudlow, C. et al. UK biobank: an open access resource for identifying the causes of a wide range of complex diseases of middle and old age. PLoS Med. 12, e1001779 (2015).

62. Bycroft, C. et al. The UK Biobank resource with deep phenotyping and genomic data. Nature 562, 203–209 (2018).

63. Chang, C. C. et al. Second-generation PLINK: rising to the challenge of larger and richer datasets. GigaScience 4, 7 (2015).

64. McCarthy, S. et al. A reference panel of 64,976 haplotypes for genotype imputation. Nat. Genet. 48, 1279–1283 (2016).

65. Loh, P.-R. et al. Reference-based phasing using the Haplotype Reference Consortium panel. Nat. Genet. 48, 1443–1448 (2016).

66. Das, S. et al. Next-generation genotype imputation service and methods. Nat. Genet. 48, 1284–1287 (2016).

67. Taliun, D. et al. Sequencing of 53,831 diverse genomes from the NHLBI TOPMed Program. Nature 590, 290–299 (2021).

68. Frankish, A. et al. GENCODE reference annotation for the human and mouse genomes. Nucleic Acids Res. 47, D766–D773 (2019).

69. Dobin, A. et al. STAR: ultrafast universal RNA-seq aligner. Bioinformatics 29, 15–21 (2013).

70. Alvarez, M. et al. Enhancing droplet-based single-nucleus RNA-seq resolution using the semi-supervised machine learning classifier DIEM. Sci. Rep. 10, 11019 (2020).

71. Hao, Y. et al. Integrated analysis of multimodal single-cell data. Cell 184, 3573–3587.e29 (2021).

72. Yang, S. et al. Decontamination of ambient RNA in single-cell RNA-seq with DecontX. Genome Biol. 21, 57 (2020).

73. Kang, H. M. et al. Multiplexed droplet single-cell RNA-sequencing using natural genetic variation. Nat. Biotechnol. 36, 89–94 (2018).

74. McGinnis, C. S., Murrow, L. M. & Gartner, Z. J. DoubletFinder: Doublet Detection in Single-Cell RNA Sequencing Data Using Artificial Nearest Neighbors. Cell Syst. 8, 329–337.e4 (2019).

75. Korsunsky, I. et al. Fast, sensitive and accurate integration of single-cell data with Harmony. Nat. Methods 16, 1289–1296 (2019).

76. Aran, D. et al. Reference-based analysis of lung single-cell sequencing reveals a transitional profibrotic macrophage. Nat. Immunol. 20, 163–172 (2019).

77. Melms, J. C. et al. A molecular single-cell lung atlas of lethal COVID-19. Nature 595, 114– 119 (2021).

78. Lake, B. B. et al. An atlas of healthy and injured cell states and niches in the human kidney. Nature 619, 585–594 (2023).

79. Ashburner, M. et al. Gene ontology: tool for the unification of biology. The Gene Ontology Consortium. Nat. Genet. 25, 25–29 (2000).

80. Liao, Y., Wang, J., Jaehnig, E. J., Shi, Z. & Zhang, B. WebGestalt 2019: gene set analysis toolkit with revamped UIs and APIs. Nucleic Acids Res. 47, W199–W205 (2019).

81. Ritchie, M. E. et al. limma powers differential expression analyses for RNA-sequencing and microarray studies. Nucleic Acids Res. 43, e47–e47 (2015).

82. Law, C. W., Chen, Y., Shi, W. & Smyth, G. K. voom: precision weights unlock linear model analysis tools for RNA-seq read counts. Genome Biol. 15, R29 (2014).

83. Langfelder, P., Luo, R., Oldham, M. C. & Horvath, S. Is my network module preserved and reproducible? PLoS Comput. Biol. 7, e1001057 (2011).

84. Kurki, M. I. et al. FinnGen provides genetic insights from a well-phenotyped isolated population. Nature 613, 508–518 (2023).

85. Corces, M. R. et al. An improved ATAC-seq protocol reduces background and enables interrogation of frozen tissues. Nat. Methods 14, 959–962 (2017).

86. Garske, K. M. et al. Reverse gene-environment interaction approach to identify variants influencing body-mass index in humans. Nat. Metab. 1, 630–642 (2019).

87. Finucane, H. K. et al. Heritability enrichment of specifically expressed genes identifies disease-relevant tissues and cell types. Nat. Genet. 50, 621–629 (2018).

88. Pulit, S. L. et al. Meta-analysis of genome-wide association studies for body fat distribution in 694 649 individuals of European ancestry. Hum. Mol. Genet. 28, 166–174 (2019).

89. Robinson, M. D. & Oshlack, A. A scaling normalization method for differential expression analysis of RNA-seq data. Genome Biol. 11, R25 (2010).

90. Buja, A. & Eyuboglu, N. Remarks on Parallel Analysis. Multivar. Behav. Res. 27, 509–540 (1992).

91. Zhou, H. J., Li, L., Li, Y., Li, W. & Li, J. J. PCA outperforms popular hidden variable inference methods for molecular QTL mapping. Genome Biol. 23, 210 (2022).

92. Shabalin, A. A. Matrix eQTL: ultra fast eQTL analysis via large matrix operations. Bioinformatics 28, 1353–1358 (2012).

93. Urbut, S. M., Wang, G., Carbonetto, P. & Stephens, M. Flexible statistical methods for estimating and testing effects in genomic studies with multiple conditions. Nat. Genet. 51, 187–195 (2019).

94. Choi, S. W., Mak, T. S.-H. & O’Reilly, P. F. Tutorial: a guide to performing polygenic risk score analyses. Nat. Protoc. 15, 2759–2772 (2020).

95. Manichaikul, A. et al. Robust relationship inference in genome-wide association studies. Bioinforma. Oxf. Engl. 26, 2867–2873 (2010).

96. Choi, S. W. & O’Reilly, P. F. PRSice-2: Polygenic Risk Score software for biobank-scale data. GigaScience 8, giz082 (2019).

97. Choi, S. W. et al. PRSet: Pathway-based polygenic risk score analyses and software. PLoS Genet. 19, e1010624 (2023).

## Supplementary References

1. Emont, M. P. et al. A single-cell atlas of human and mouse white adipose tissue. Nature 603, 926–933 (2022).

2. Morabito, S., Reese, F., Rahimzadeh, N., Miyoshi, E. & Swarup, V. hdWGCNA identifies co-expression networks in high-dimensional transcriptomics data. Cell Rep. Methods 3, 100498 (2023).

